# Deep-learning-based Prediction of Late Age-Related Macular Degeneration Progression

**DOI:** 10.1101/19006171

**Authors:** Qi Yan, Daniel E. Weeks, Hongyi Xin, Anand Swaroop, Emily Y. Chew, Heng Huang, Ying Ding, Wei Chen

**Author notes:** Correspondence and requests for materials should be addressed to: Q.Y., Y.D., or W.C. These authors jointly supervised this work (Qi Yan, Ying Ding, Wei Chen).

## Abstract

Both genetic and environmental factors influence the etiology of age-related macular degeneration (AMD), a leading cause of blindness. AMD severity is primarily measured by fundus images and recently developed machine learning methods can successfully predict AMD progression using image data. However, none of these methods have utilized both genetic and image data for predicting AMD progression. Here we jointly used genotypes and fundus images to predict an eye as having progressed to late AMD with a modified deep convolutional neural network (CNN). In total, we used 31,262 fundus images and 52 AMD-associated genetic variants from 1,351 subjects from the Age-Related Eye Disease Study (AREDS) with disease severity phenotypes and fundus images available at baseline and follow-up visits over a period of 12 years. Our results showed that fundus images coupled with genotypes could predict late AMD progression with an averaged area under the curve (AUC) value of 0.85 (95%CI: 0.83-0.86). The results using fundus images alone showed an averaged AUC of 0.81 (95%CI: 0.80-0.83). We implemented our model in a cloud-based application for individual risk assessment.

## INTRODUCTION

Age-related macular degeneration (AMD) is the leading cause of blindness among older adults in Caucasians ^1-3^. It is a progressive neurodegenerative disease influenced by both environmental and genetic risk factors ^4-6^. AMD severity is mainly diagnosed by color fundus images in a clinical setting by ophthalmologists. Late AMD comes in two forms: (1) Geographic atrophy (GA) also known as dry AMD is characterized by a gradual degeneration and disappearance of retinal pigment epithelium, photoreceptor cells, and the choriocapillaris layer in the central retina; (2) choroidal neovascularization (CNV) also known as wet AMD is characterized by the growth of new, leaky blood vessels into the retina causing widespread photoreceptor loss and ultimately rapid decline in visual acuity ^7^. Some patients with early or intermediate stage AMD maintain their vision for a long time with slow disease progression over time, but others quickly progress to one or both forms of late AMD.

Genetics plays a critical role in AMD pathogenesis. Genome-wide association studies (GWAS) and sequencing studies have identified many variants that are associated with AMD ^8-11^. For example, a total of 52 independent genetic markers including both common and rare variants from 34 loci were reported to have associations with AMD risk in a recent large-scale genome-wide association study (GWAS) by the International AMD Genomics Consortium ^9^. In addition to the successes in identifying AMD-related genetic markers from case-control designs, a recent study of AMD progression risk using the Age-Related Eye Disease Study (AREDS) dataset ^12^ showed that some of the known AMD risk variants could also influence progression time to late AMD ^10^.

In parallel to genetic studies, machine learning methods, particularly deep convolutional neural networks (DCNN) have been useful in image recognition and classification in ophthalmology. CNN have been used for the aforementioned automated AMD grading, identifying diabetic retinopathy and cardiovascular risk factors from fundus images, and interpreting and segmenting optical coherence tomography (OCT) images ^13-20^. As opposed to traditional machine learning approaches that rely on “feature engineering”, which involves computing features explicitly defined by experts ^21-23^, CNN can learn features directly from the images themselves. CNN is a family of deep learning techniques characterized by enabling the networks to contain many computation layers that can automatically, deeply and comprehensively learn features from lower-level structures to more generalized higher-level structures. Recently several studies have used the color fundus images to perform automated AMD grading ^17,18,20^ and estimation of future risk of AMD ^24^ by applying convolutional deep learning methods. However, none of these methods consider genetic data in the prediction model.

In addition to using fundus images for AMD grading, in conjunction with genotypes, fundus image data can be used to predict the probability of late AMD progression exceeding certain inquired durations. Since late AMD is irreversible, such prediction could urge potential patients to start preventative care beforehand and slow down the disease progression. The Age-Related Eye Disease Study (AREDS), a large-scale clinical trial from the National Eye Institute, includes massive genome-wide genotyping data, longitudinal color fundus photographs, and disease severity assessment over a period of 12 years, providing an unprecedented opportunity for us to investigate AMD progressing using both dynamic (fundus image) and static (genetics) information.

In this study, we jointly used genotypes and fundus images to predict an eye as having progressed to late AMD (which may never occur) within certain inquired durations from the current visit. The inquired duration was selected in advance, and it was relative to the time when the fundus image was taken, not to the time of the baseline visit. Specifically, for one eye, the inputs included one fundus image taken at the current visit and the genotypes of the corresponding subject, and the output was the probability that the time to late AMD exceeds the inquired duration. To our knowledge, this is the first time to jointly use genotypes and fundus images in a prediction model for AMD risk and progression.

## RESULTS

### Study Data Characteristics

Of the AREDS participants, 1,351 Caucasians who had at least one eye free of late AMD at baseline and at least one follow-up visit had all information on images and genotypes (Table 1). The baseline mean age was 68.8 (SD=5.0) years. About 56% (N=750) of participants were females. About 46% were never smokers (N=626), another 47% were former smokers (N=634) and 7% were current smokers (N=91). Smoking status was defined at the baseline visit. The participants had mean follow-up time of 10.3 (SD=1.6) years and they were followed up every 6 months in the first 6 years to every 1 year after year 6. 2,678 eyes of the 1,351 participants were not in the late AMD stage at baseline. These eyes had a low mean severity score at baseline of 3.9 (SD=3.2), because the majority of eyes had low baseline severity scores (54% eyes with baseline severity score 1-3, 23% eyes with 4-6 and 24% eyes with 7-8). Moreover, only 4% eyes with baseline severity 1-3 progressed to late AMD by the end of the follow-up time, 50% eyes with 4-6 progressed by the end of the follow-up time, and 92% eyes with 7-8 progressed by the end of the follow-up time. In addition, the number of useable fundus images (i.e., the fundus images of each eye at each visit with corresponding genotypes) for prediction decreased as the progression inquired year increased from 2 to 7 years due to the censored subjects (Table 1). The inquired years were defined as well as an illustrative example was provided in the Methods section. The number of useable fundus images for predicting whether the progression time to late AMD exceeded the inquired years were shown in Supplementary Figure 1. Note that the useable fundus images included the ones at both baseline and the follow-up visits.

**Table 1.** Characteristics of the participants

|  | **AREDS** | **Training** | **Test** |
| --- | --- | --- | --- |
| Subject-level variables | 1,351 subjects | 1,223 subjects | 128 subjects |
| Baseline age, year (mean ± SD) | 68.8 ± 5.0 | 68.8 ± 5.0 | 68.5 ± 4.8 |
| Female (N, %) | 750 (55.5) | 682 (55.8) | 68 (53.1) |
| Follow-up time, (mean ± SD) | 10.3 ± 1.6 | 10.2 ± 1.7 | 10.9 ± 1.0 |
| Baseline smoking status (N, %) |  |  |  |
| Never smoked | 626 (46.3) | 566 (46.3) | 60 (46.9) |
| Former smoker | 634 (46.9) | 576 (47.1) | 58 (45.3) |
| Current smoker | 91 (6.7) | 81 (6.6) | 10 (7.8) |
| Eye-level variables | 2,678 eyes | 2,422 eyes | 256 eyes |
| Baseline AMD severity score at eye-level |  |  |  |
| Mean ± SD | 3.9 ± 3.2 | 4.0 ± 3.2 | 3.9 ± 3.2 |
| 1-3 (n, %) | 1,442 (53.8) | 1,310 (54.1) | 132 (51.6) |
| 4-6 (n, %) | 600 (22.5) | 528 (21.8) | 72 (28.1) |
| 7-8 (n, %) | 636 (23.7) | 584 (24.1) | 52 (20.3) |
| Progressed eyes with baseline severity |  |  |  |
| 1-3 (n, %) | 50 (3.5) | 48 (3.7) | 2 (1.5) |
| 4-6 (n, %) | 300 (50.0) | 260 (49.2) | 40 (55.6) |
| 7-8 (n, %) | 585 (92.0) | 537 (92.0) | 48 (92.3) |
| Observation-level variables |  |  |  |
| Fundus images used for prediction with progression cutoff |  |  |  |
| 2 years (n) | 27,499 | 24,654 | 2,845 |
| 3 years (n) | 25,862 | 23,170 | 2,692 |
| 4 years (n) | 24,287 | 21,709 | 2,578 |
| 5 years (n) | 22,435 | 20,041 | 2,394 |
| 6 years (n) | 20,240 | 18,118 | 2,122 |
| 7 years (n) | 18,066 | 16,172 | 1,894 |

### Predicting Progression Time to Late AMD Exceeding Inquired Years Using Fundus Images Alone

First, we tested the ability of our proposed network to predict the progression time to late AMD exceeding inquired years using the AREDS fundus images alone (Supplementary Figure 2a). The partitioning of training and testing datasets and the selection of late AMD progression inquired years were previously explained. The ROC and AUC (Figure 1) and Brier scores on the testing dataset are reported in Table 2. The performance of our CNN showed promising results even when using fundus images alone. In the testing dataset, the model achieved AUC range from 0.79 (95% CI [confidence interval]: 0.78-0.81) to 0.84 (95% CI: 0.82-0.86) for progression inquired year 2 ∼ 7 (Figure 1 and Table 2). Furthermore, in order to improve the interpretability of our model, we added a secondary output layer for current AMD severity between the final convolutional layer and the primary output for the progression time to late AMD (Supplementary Figure 2b). The model achieved similar AUC range between 0.79 (95% CI: 0.77-0.81) and 0.84 (95% CI: 0.83-0.86) for progression inquired year 2 ∼ 7 (Figure 1 and Table 2). Besides, the model automatically graded the AMD severity based on fundus images with an accuracy range of 0.56 to 0.60 for progression inquired year 2 ∼ 7 (Supplementary Table 1). Note that the random accuracy is 0.33. The density curves of predicted probability of having late AMD progression time before or after each of the six inquired years were generated (Supplementary Figure 3 [the first and third columns]) to visually examine the accuracy of before and after inquired year prediction separately. The results showed that CNN could accurately predict the probability of having late AMD progression time exceeding the inquired year. However, although most of the eyes with progression time before the inquired year could be correctly predicted when Youden indices ^25^ (Supplementary Table 2) were used as the thresholds to dichotomize the predictions, a sizeable number of eyes were falsely predicted as having a progression time exceeding the inquired year.

**Table 2.**
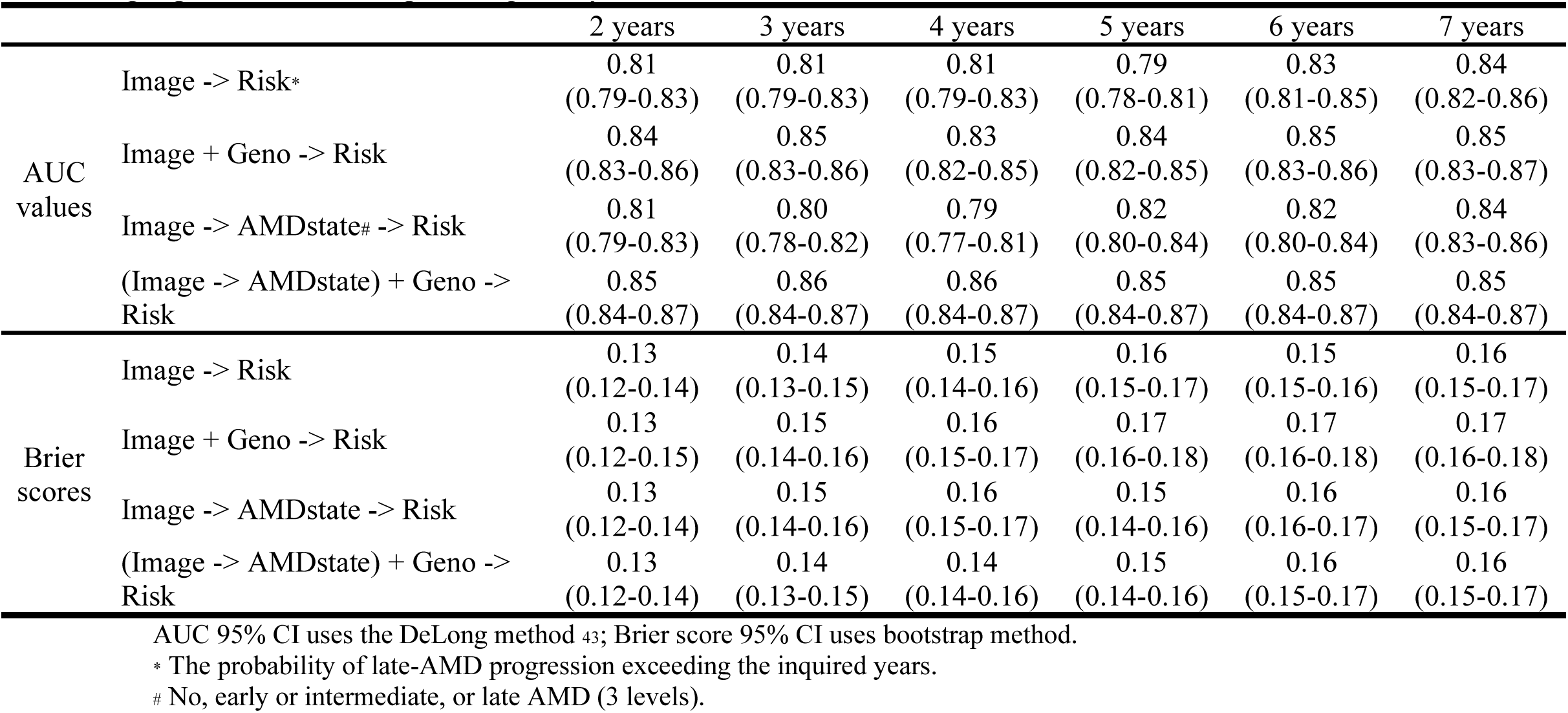
AUC values (95% CI) and Brier scores (95% CI) of the prediction of probability of late-AMD progression exceeding the inquired years for four models

|  |  | 2 years | 3 years | 4 years | 5 years | 6 years | 7 years |
| --- | --- | --- | --- | --- | --- | --- | --- |
| AUC values | Image -> Risk* | 0.81<br>(0.79-0.83) | 0.81<br>(0.79-0.83) | 0.81<br>(0.79-0.83) | 0.79<br>(0.78-0.81) | 0.83<br>(0.81-0.85) | 0.84<br>(0.82-0.86) |
|  | Image + Geno -> Risk | 0.84<br>(0.83-0.86) | 0.85<br>(0.83-0.86) | 0.83<br>(0.82-0.85) | 0.84<br>(0.82-0.85) | 0.85<br>(0.83-0.86) | 0.85<br>(0.83-0.87) |
|  | Image -> AMDstate# -> Risk | 0.81<br>(0.79-0.83) | 0.80<br>(0.78-0.82) | 0.79<br>(0.77-0.81) | 0.82<br>(0.80-0.84) | 0.82<br>(0.80-0.84) | 0.84<br>(0.83-0.86) |
|  | (Image -> AMDstate) + Geno -> Risk | 0.85<br>(0.84-0.87) | 0.86<br>(0.84-0.87) | 0.86<br>(0.84-0.87) | 0.85<br>(0.84-0.87) | 0.85<br>(0.84-0.87) | 0.85<br>(0.84-0.87) |
| Brier scores | Image -> Risk | 0.13<br>(0.12-0.14) | 0.14<br>(0.13-0.15) | 0.15<br>(0.14-0.16) | 0.16<br>(0.15-0.17) | 0.15<br>(0.15-0.16) | 0.16<br>(0.15-0.17) |
|  | Image + Geno -> Risk | 0.13<br>(0.12-0.15) | 0.15<br>(0.14-0.16) | 0.16<br>(0.15-0.17) | 0.17<br>(0.16-0.18) | 0.17<br>(0.16-0.18) | 0.17<br>(0.16-0.18) |
|  | Image -> AMDstate -> Risk | 0.13<br>(0.12-0.14) | 0.15<br>(0.14-0.16) | 0.16<br>(0.15-0.17) | 0.15<br>(0.14-0.16) | 0.16<br>(0.16-0.17) | 0.16<br>(0.15-0.17) |
|  | (Image -> AMDstate) + Geno -> Risk | 0.13<br>(0.12-0.14) | 0.14<br>(0.13-0.15) | 0.14<br>(0.14-0.16) | 0.15<br>(0.14-0.16) | 0.16<br>(0.15-0.17) | 0.16<br>(0.15-0.17) |

AUC 95% CI uses the DeLong method <sup>43</sup>; Brier score 95% CI uses bootstrap method.

**Figure 1.**
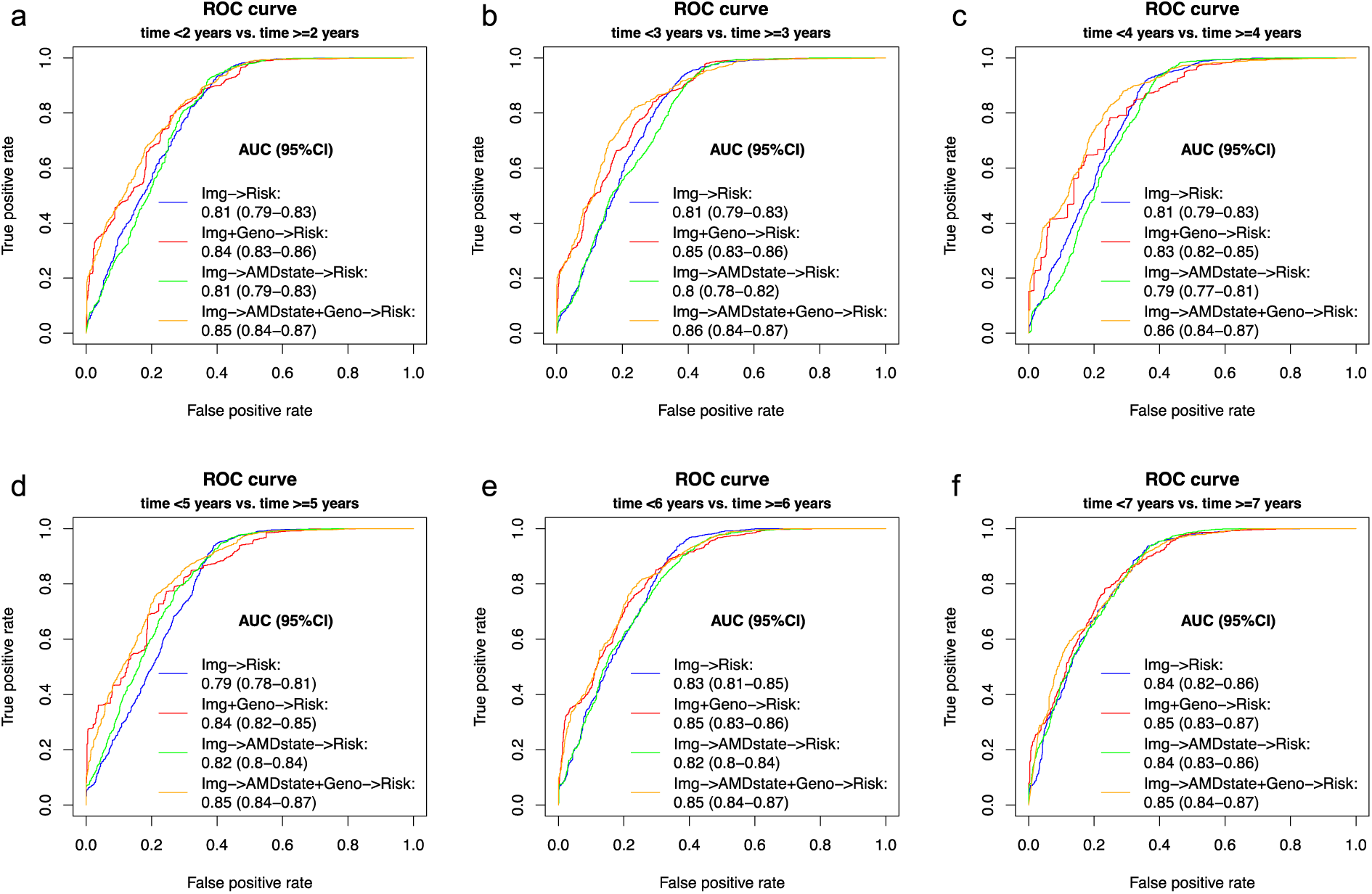
ROC curves of the prediction of late AMD progression time exceeding the inquired years for four models. **The four models are** (1) fundus images predicting late-AMD progression exceeding the inquired years; (2) fundus images + genotypes predicting late-AMD progression exceeding the inquired years; (3) fundus images both classifying current AMD severity and predicting late-AMD progression exceeding the inquired years; and (4) fundus images + genotypes both classifying current AMD severity and predicting late-AMD progression exceeding the inquired years. (a-f) inquired years from 2 to 7.

### Model Interpretation for Supporting Clinical Decision

Next, we generated saliency maps to visualize the important regions that had the greatest impact on the model predictions using the fundus images only models. Generally, the CNN should be able to detect the macula region and make decisions based on the features (e.g., drusen) in this region. Three representative subjects of left eye fundus images with accompanying saliency maps for each inquired year prediction were shown in Figures 2-4. Subject #1 (Figure 2) had 4 visits and progressed to late AMD at year 4.8; Subject #2 (Figure 3) had 4 visits and did not progress to late AMD by the end of 11.1-year follow-up; and Subject #3 (Figure 4) had 5 visits and was at late AMD status at baseline. Thus, Subject #1 who progressed in the middle of the follow-up was more challenging to predict than Subject #2 who did not progress and Subject #3 who progressed before baseline time. Figure 2 showed that Subject #1 had early/intermediate AMD at the first three visits and was labeled as late AMD at the fourth visit. Most models with different cut-off years gave accurate predictions. However, the model with inquired year equal to 3 tended to predict a long progression time when small drusen was observed. This was also the reason that the density curves with only images as predictors led to a second peak on the right side (blue curves in Supplementary Figure 3). In other words, the samples that were falsely predicted to the category exceeding the inquired years were true labeled in the category before the inquired years. The results showed that genotypes could help to correct this misclassification. Figure 3 showed that Subject #2 had healthy macula at all visits and the models correctly predicted that the subject had a long progression time. Please note that the true labels were missing at visit year 5.8 for inquired years equal to 6 and 7, because this subject was censored and had 5.3 more follow-up years at visit year 5.8. Thus, at visit year 5.8, this subject could finally progress to late AMD before or after 6 more years, which was ambiguous to use this cutoff, same to the cutoff of year 7. Figure 4 showed that Subject #3 had late AMD since baseline, and all models could detect the drusen and correctly assign it to the right category.

**Figure 2.**
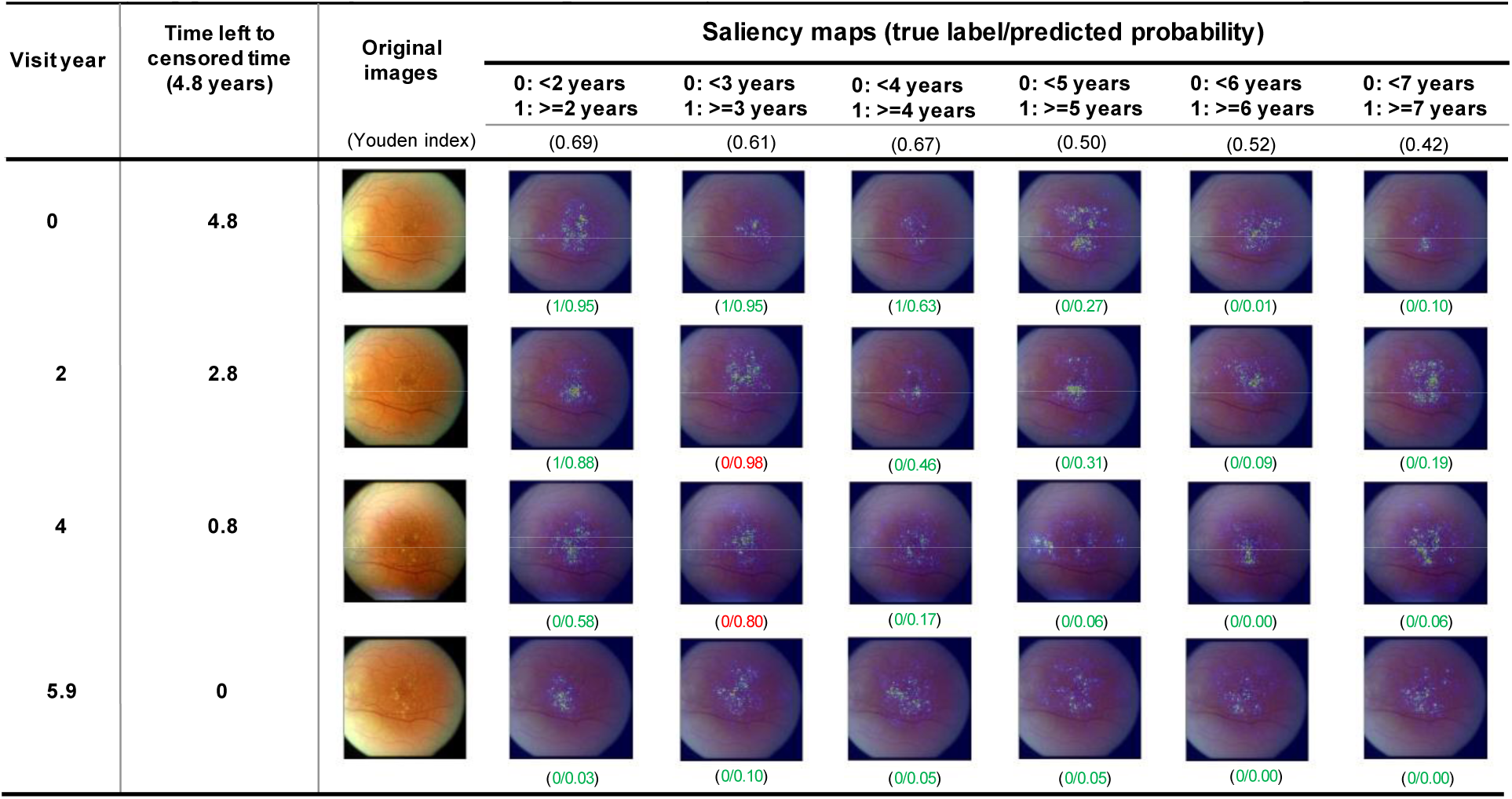
Saliency maps for left eye of Subject #1 over 5.9 years. This subject progressed to late AMD after 4.8 years of follow-up. The highlighted dots indicate the area that the CNN learned to make the decision. The first number in the parenthesis is the true label (1=not progressed, 0=progressed) and the second number is the estimated probability of late AMD progression time exceeding the inquired year relative to the current visit. The green numbers indicate accurate predictions and red numbers indicate inaccurate predictions using Youden indices (Supplementary Table 2, Img -> Risk) as the thresholds to dichotomize the predictions.

**Figure 3.**
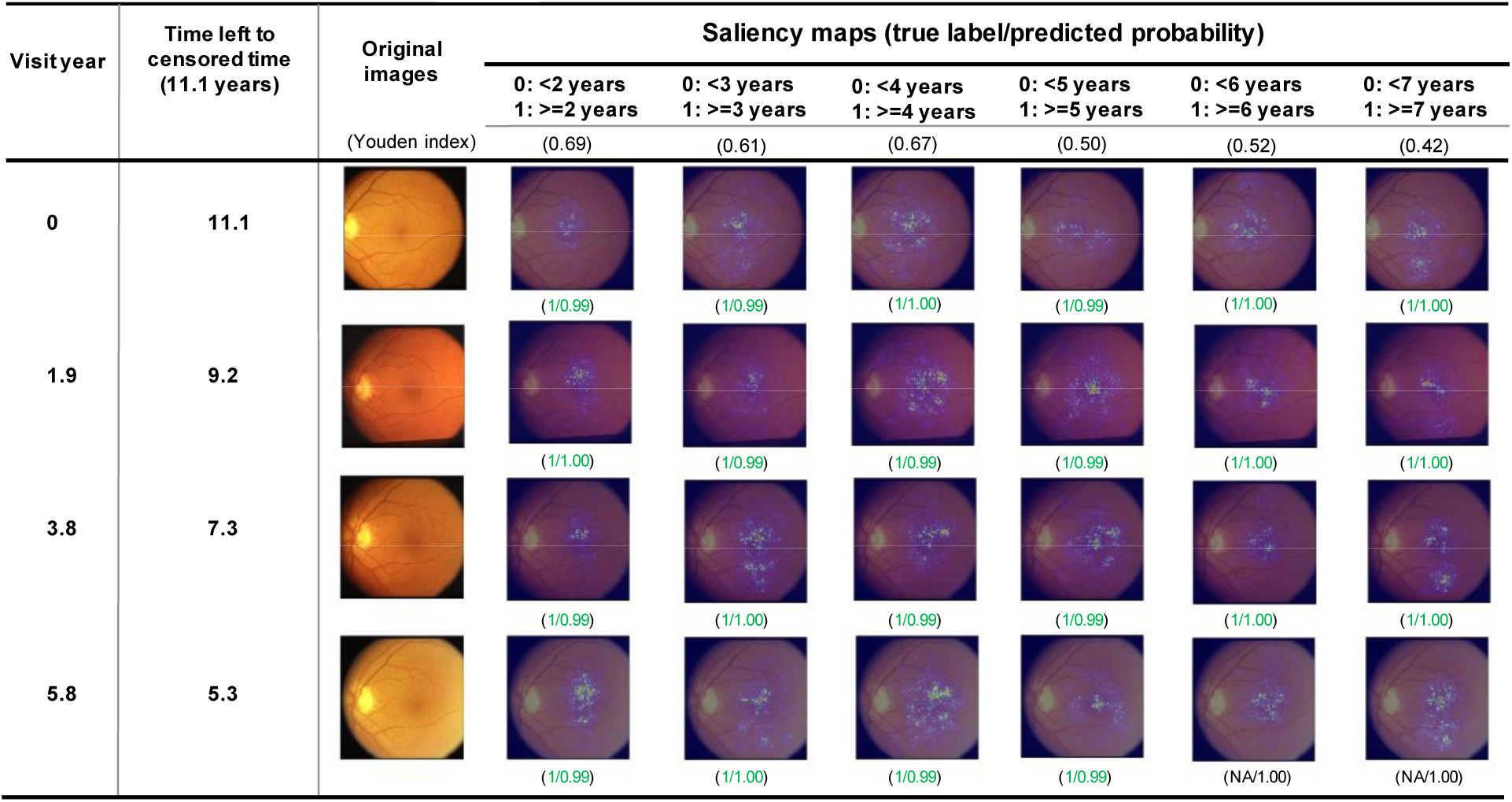
Saliency maps for left eye of Subject #2 over the first 5.8 years. This subject was censored after 11.1 years of follow-up. The highlighted dots indicate the area that the CNN learned to make the decision. The first number in the parenthesis is the true label (1=not progressed, 0=progressed, NA=progression status unknown) and the second number is the estimated probability of late AMD progression time exceeding the inquired year relative to the current visit. The green numbers indicate accurate predictions and red numbers indicate inaccurate predictions using Youden indices (Supplementary Table 2, Img -> Risk) as the thresholds to dichotomize the predictions.

**Figure 4.**
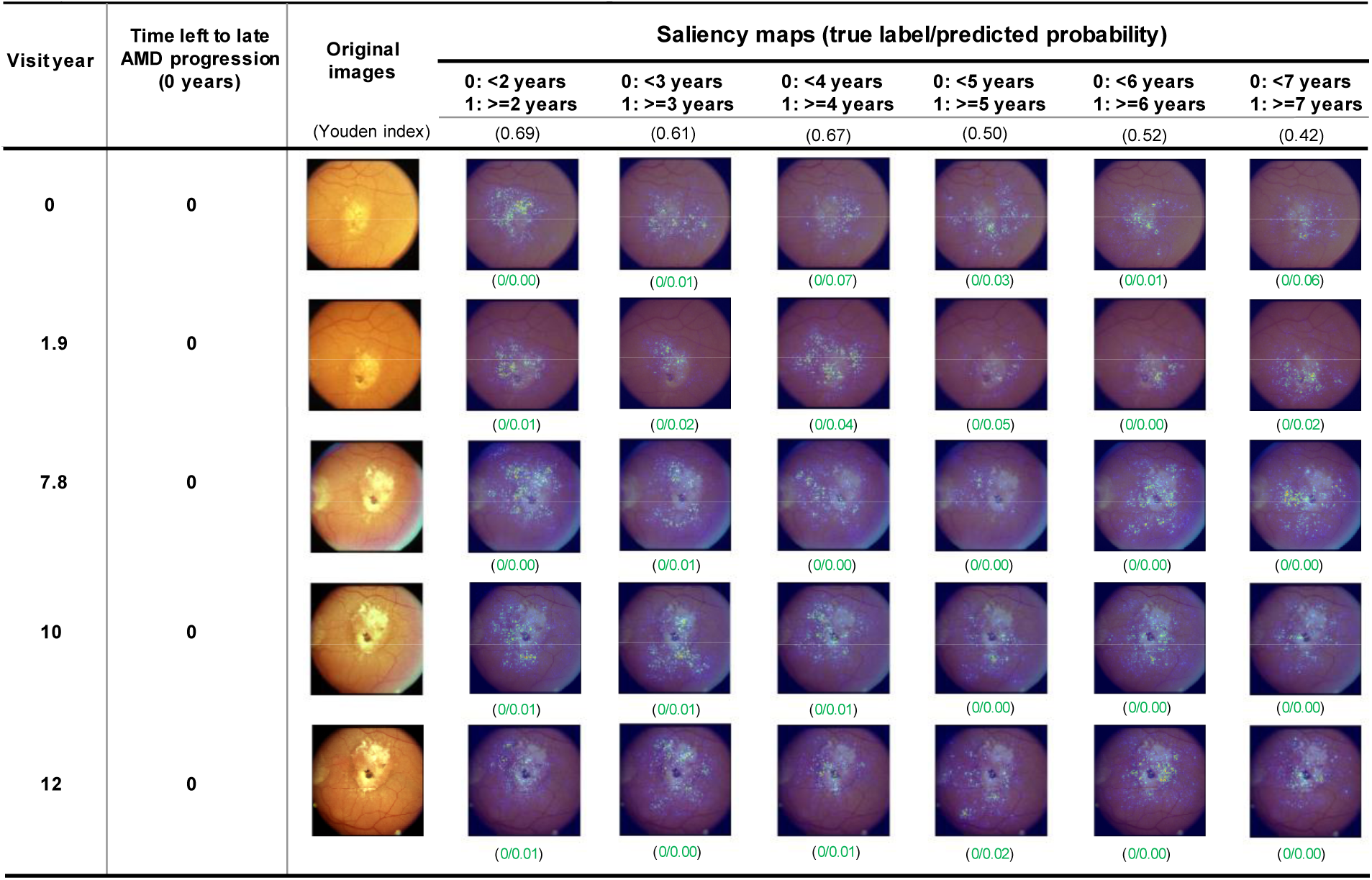
Saliency maps for left eye of Subject #3 over 12 years. This subject developed late AMD before enrollment. The highlighted dots indicate the area that the CNN learned to make the decision. The first number in the parenthesis is the true label (1=not progressed, 0=progressed) and the second number is the estimated probability of late AMD progression time exceeding the inquired year relative to the current visit. The green numbers indicate accurate predictions and red numbers indicate inaccurate predictions using Youden indices (Supplementary Table 2, Img -> Risk) as the thresholds to dichotomize the predictions.

### Predicting Progression Time to Late AMD Exceeding Inquired Years Using Fundus Images along with genotypes

In addition to fundus images, we added 52 AMD associated independent genetic variants reported by the International AMD Genomics Consortium ^9^ to the model (Supplementary Figure 2c). Similarly, we tested the scenarios with and without AMD severity as a secondary output (Supplementary Figure 2d). When evaluating the performance using the test dataset, the models with AMD severity as a secondary output achieved slightly higher AUC (range between 0.85 [95% CI: 0.84-0.87] and 0.86 [95% CI: 0.84-0.87] for progression inquired year 2 ∼ 7) than without AMD severity (range between 0.83 [95% CI: 0.82-0.85] and 0.85 [95% CI: 0.83-0.86], Figure 1 and Table 2). The accuracy for AMD severity grading was from 0.57 to 0.60 (Supplementary Table 1). The AMD severity as a secondary output helped to better explain the prediction model that the fundus images could be used for automated AMD severity grading, and then the AMD severity at the current visit could predict late AMD progression exceeding the inquired years. Without AMD severity, we only knew fundus images could predict late AMD progression exceeding the inquired years, but missed the related features (i.e., current AMD severity) in between. These results also indicated that genetics could noticeably improve the AUCs for AMD progression time (Table 2). The density curves of predicted probability of having AMD progression time before or after the six inquired years (Supplementary Figure 3) showed that the addition of genetics in the neural network could correct the falsely predicted eyes from the group of after the inquired year to the correct group of before the inquired year. This is evidenced by that the blue curves on the right (Supplementary Figure 3) were much lower in models with genetics than models without genetics.

The feature importance heatmaps (Supplementary Figures 4-6) from LIME for the sub-network using fundus images along with genotypes (Supplementary Figure 7b inside the red rectangle) to predict late AMD progression time for subjects #1, #2 and #3 showed the first two variables contributing the most to the predictions for each visit year. Although the summarized image inputs (a size of 2,048) and the SNP inputs (a size of 52) were unbalanced, SNPs were always shown among the most important variables (*e.g*., the SNP from *ARMS2/HTRA1* appears in 23 of 24 models).

### Replication Study Using Independent UK Biobank Dataset

In addition to the test dataset we generated from dbGaP, we extracted a set of 200 Caucasian subjects from the UK Biobank^26^ as an independent test dataset, and used 3 years as the inquired year. The model (4) showed an AUC of 0.9 (95%CI: 0.85-0.94) for predicting whether the eye progresses to late AMD exceeding the 3 years (Supplementary Figure 8). Again, please note that we treated the AMD patients from UK Biobank as late AMD patients, although some of them could be at early/intermediate stages.

## DISCUSSION

Our results show that the application of CNN to retinal fundus images coupled with genotypes can be used to predict the probability of late AMD progression exceeding the inquired years and diagnose the current AMD severity. The results indicate that the fundus images alone can predict late AMD progression exceeding certain inquired years with reasonable accuracy. The addition of secondary inputs of genotypes can noticeably improve accuracy (Table 2). The addition of a secondary output of AMD severity further improve the model interpretability. We also conducted an additional model using current age and baseline smoking status as predictors in addition to fundus images and genetics (Supplementary Figure 9), but they did not improve the prediction performance, probably because the age and smoking effects were already reflected in fundus images or the dynamic age range for those who progressed to late-AMD in our training cohort is limited across the range of AMD pathology. Although the training of a deep learning model is computationally intensive, once the model is trained and weights are saved, the prediction of a new subject takes only a few seconds.

The goal of our study is different from previous ones, which mainly used fundus images for automated grading of AMD severity scores. Since the severity score is graded by ophthalmologists merely based on the fundus images, those studies did not engage other phenotypes or genotypes, although it is known that AMD is associated with age, smoking status and a number of genetic variants. We believe that the prediction of late AMD progression time is more useful as compared to the current severity score for patients to start preventative care early. Furthermore, the automated classification using directly measured variables would also help reduce the discrepancy among human graders and reduce costs of large-scale image assessment projects.

We used the pre-designed CNN architecture and pre-trained weights as our initial values, which are used for general image classification of thousands of objects based on millions of images. For our specific fundus image task, this helps the model learn more accurately with less data, because the existing CNN architecture and trained weights can identify simple features (e.g., edges and orientation) and further combine them to more complex objects, which mimics the way our visual system works. Analogously, our proposed model could mimic the way that ophthalmologists interpret the fundus images by identifying macula, drusen, pigmentary changes, etc. The saliency maps indicate that our model pays attention to assess features in the macula region in the retina to predict late AMD progression exceeding certain inquired years, which is as expected. Furthermore, the model might capture some unknown features that are important for AMD progression, but neglected by ophthalmologists.

In addition to AUC values and Brier scores, we were able to calculate the prediction accuracy after dichotomizing the prediction risk by using the Youden indices ^25^ (Supplementary Table 2) as the cutoff, which is based on the AUC curve. Our results showed that the fundus images coupled with genotypes could predict late AMD progression with an averaged accuracy (SD) of 78.4% (1.6%). The results using fundus images alone showed a similar averaged accuracy (SD) of 80.0% (2.7%). The AREDS fundus images have been used in several prediction studies. Although their outcomes were different from ours, it is worth checking their accuracies. One study ^17^ classifying 13 classes (9 AREDS steps, 3 late AMD stages, and 1 for ungradable images) had an overall accuracy of 63.3%. The second study ^18^ performing two-class classification to distinguish the disease-free/early stages from the referable intermediate/late stages yielded accuracy that ranged between 88.4% and 91.6%. The third study ^20^ classifying the AREDS Simplified Severity Scale (score 0-5) had an accuracy of 67.1%. In addition to classifying the current AMD status, one study ^24^ estimating 5-year risk of AMD achieved accuracy of 75.7% for the 4-step and 59.1% for the 9-step AMD severity scales. Moreover, another study ^27^ using optical coherence tomography (OCT) images to predict eyes with intermediate AMD progressing to CNV or GA yield AUC of 0.68 and 0.80 for CNV and GA respectively.

Here we included only 52 AMD risk-associated SNPs in our prediction models. To evaluate the effect of including a larger number of SNPs, we conducted another set of analyses by using the SNPs with *P*-value < 1×10^−5^ from a GWAS of AMD progression ^10^ (n = 2,721 subjects). In total, 1,057 SNPs were included and we used the cutoff of year 3 for illustration. Please note that the previous 52 SNPs were identified from a large GWAS of AMD case-control study ^9^ (43,566 subjects). Although AMD progression is a better phenotype than AMD case-control status, the sample size of the AMD progression GWAS is much smaller. Finally, 7 out of the previous 52 SNPs were also on the new list of 1,057 SNPs. The results (Supplementary Figure 10) showed that AUC = 0.84 for Img+Geno−>Risk and 0.84 for Img−>AMDstate+Geno−>Risk. On the other hand, the 52 SNP results showed that AUC = 0.85 for Img+Geno−>Risk and 0.86 for Img−>AMDstate+Geno−>Risk. Thus, the 52 SNPs identified from a much larger study of AMD risk with *P*-value < 5×10^−8^, although the phenotype is not AMD progression, were slightly better than 1,057 SNPs identified from a relatively small AMD progression study with *P*-value < 1×10^−5^. Even 1,057 is not a very large number and can be handled by a fully connected neural network (NN), however, if the genome-wide SNPs (e.g., several millions) are used as the input, the fully connected NN is not expected to work in terms of computational intensity. In this case, some other alternative solutions might be considered: 1. using a CNN for SNPs too; or 2. using a network reflecting the SNP-gene-pathway hierarchical structure that is similar to the idea of Hao et al ^28^. In other words, after the input layer of SNPs, only the SNPs in the same gene region are connected to a neuron in the first hidden layer, then only the genes in the same pathway are connected to a neuron in the second hidden layer that is further connected to an output layer. In this way, the number of weights needed to be estimated is much smaller than the number in a fully connected NN.

Our study has some limitations. First, this study mainly relies on the AREDS dataset. Although an independent dataset from UK Biobank is used, the available outcome is any AMD rather than late AMD. Even though there are no other large longitudinal AMD studies with both fundus images and genotypes available, it would be beneficial to evaluate our models on a separately collected dataset. Second, AREDS contains a large number of normal to early AMD eyes at the beginning of the study and many of them did not progress to the late AMD by the end of the follow-up. Although we used stepwise binary predictions instead of predicting a continuous time to try to fully use the censored eyes, we still lose some of them. Third, we used stepwise binary predictions to approximate the progression process. However, since each model was trained separately, this could lead to inconsistent results from the separate models, especially for the eyes that are hard to predict. It would be ideal to directly model the survival outcome in a single model. One possible way is to replace the final loss function with a survival type likelihood, such as the Cox partial likelihood. We plan to explore this direction in a future work. Another potential limitation is that only fundus images were used in this study. It would be desirable to have a coherent prediction by using multiple types of images (e.g., optical coherence tomography and fundus autofluorescence images). In addition, the fundus images in AREDS were collected on both eyes over several years from the same participants. In the current study, the covariance between two eyes or between different visits was not considered. Although it is hard to consider the information of correlated images using deep learning approaches, incorporating such covariance in the model could increase the prediction accuracy, which might be implemented by modifying the loss function at the output layer from the neural network. In this study, the complex neural network might implicitly capture this information that different eyes and visits from subjects having the identical genotypes could be from the same subject. To evaluate the impact of adding the status of the other fellow eye, we conducted an additional analysis. Instead of directly modeling the correlation between two eyes, we added the other eye’s current AMD severity scale as an additional input for the study eye’s prediction, although the other eye’s current AMD severity should be unknown, because our models were designed for only fundus images and genotypes available and predicting current AMD severity and progression time without the need of image specialists. Here, we just used this analysis to evaluate the impact of adding the status of the other eye. The results showed that adding the status of the other eye could improve the prediction accuracy of the study eye to an AUC of 0.91 (Supplementary Figure 11). In the future direction, a bivariate approach that simultaneously models the dependency between the two eyes, as well as to provide each eye-level and joint subject-level progression probabilities is worthwhile to pursue.

In summary, this study showed that deep learning approaches could be used to automatically predict late AMD progression exceeding certain inquired years and classify the current AMD severity stages. The joint use of fundus images and genotypes can achieve good prediction accuracy. The deep learning methods could serve an important role in decision support systems for eye services by reducing assessment time, workload and financial burden by automated analysis. Such automated analysis identifying individuals who should be referred to a specialist could become increasingly acceptable to both patients and ophthalmologists. We also developed a web-based application at http://www.pitt.edu/~qiy17/amdprediction.html. To our knowledge, this is the first cloud-based prediction website for AMD with deep learning techniques. In addition to distinguishing retinal pathologies using the fundus images, such as AMD, this study can be extended to be applicable to other diseases associated with images, genotypes and phenotypes (e.g., Alzheimer’s disease).

## METHODS

### Study Population and Phenotype Definition

The study subjects were from the AREDS study sponsored by the National Eye Institute ^12^, which was a long-term, multicenter, prospective study of AMD and age-related cataracts with a 12-year follow-up period to assess the risk factors and impact of daily supplements. Eligible subjects were between 55 and 80 years old at baseline and free of sight-threatening conditions other than cataract or AMD. In this study, we confined our analyses to the AMD data and only used Caucasian participants with genotype data and at least one follow-up visit.

In the analyses, we used 31,262 color fundus images centered above the macula from 1,351 subjects with corresponding genotypes and phenotypes available at baseline and follow-up visits. The detailed image taking procedure was described elsewhere ^29^. The AREDS AMD scale ^30^, based upon severity score from 1 to 12, was adopted to determine whether the eye was in the late AMD stage or not, which was measured based on centralized grading of these fundus images obtained at each semi-annual/annual follow-up visit ^30^. The progression of early/intermediate to late AMD is often distinguished by the growth of drusen size and/or pigmentary abnormalities at the macula region ^31,32^. For each non-late AMD eye at each visit, we calculated its time period between the current visit and the time to late AMD, defined as the time to the first visit when the severity score reached 9 (noncentral GA) or higher (10: central GA, 11: CNV, and 12: CNV and central GA). If the eye’s severity score was less than 9 by the end of follow-up, the time to late AMD was treated as censored at the last visit.

### Prediction of the Probability that the Time to Late AMD Progression Exceeds an Inquired Year

Since the exact times to late AMD of censored eyes were unknown and we only knew the eyes were not progressed at certain time points, in order to make the full use of the available information, we performed a set of binary predictions instead of predicting a continuous progression time. Specifically, we predicted the probability that the time to late AMD progression exceeds the inquired durations, 2, 3, 4, 5, 6 and 7 years from the current visit. This implies some eyes cannot be used for certain of these prediction models. For example, if one subject was censored at year 5.5 without having progressed to late AMD, this subject’s eyes were used for inquired years 2, 3, 4 and 5, because it was known that the eyes had not progressed by those inquired years. However, these eyes could have progressed on either side of the inquired years of 6 and 7, which made this eye unusable for these two inquired years. In addition to the late AMD scores (i.e., 9-12), a severity score of 1 indicates little or no AMD-related changes, whereas scores 2 through 8 indicate early or intermediate AMD ^30^. Other variables we considered were current visit age and baseline smoking status (never, former, current).

### Replication dataset

In addition to AREDS, we extracted 100 Caucasian subjects labeled with AMD at baseline visit and 100 Caucasian subjects without AMD reported by the end of at least 3 years of follow-up from UK Biobank^26^ as an independent test dataset. For samples labeled with AMD, we only kept the ones with clear drusen with the help from experienced image specialist. In other words, we had 100 subjects who progressed to AMD within 3 years and 100 subjects who progressed to AMD after 3 years (it is possible that they never progressed to AMD). All the subjects needed to have genotypes and high-quality fundus images at baseline. Please note that not all of the AMD subjects had late AMD and some had early/intermediate stages of AMD. The outcome was not exactly the same as we used in the training process, although we could assume that the early/intermediate AMD subjects might progress to late AMD in a short time (e.g., within 3 years).

### Genotype Data

DNA samples from consenting subjects in AREDS were collected and genotyped centrally by the International AMD Genomics Consortium, as described previously ^9^. In brief, a custom-modified HumanCoreExome array by Illumina was used to obtain the genotypes followed by imputation with the 1000 Genomes Project reference panel (Phase I). In this study, we used 52 independent genetic variants from 34 loci that were either confirmed or newly discovered to have associations with AMD risk in a GWAS by the International AMD Genomics Consortium ^9^. We used additive genotypes (i.e., 0: no minor allele; 1: one copy of the minor allele; 2: two copies of the minor allele).

### Data Partitioning

The total of 31,262 original images were first randomly divided into a training set (90% of the subjects) and testing set (10% of the subjects). Then, the training set was further divided into 10 folds so that 10-fold cross-validation was performed with 9 folds for training and 1 fold for validation and this process was repeated 10 times. Because each subject includes multiple images over 12-year visits on both eyes, we performed this separation at the subject level, which means that images from the same subject were included in the same fold.

### Data Augmentation

A data augmentation procedure was applied to increase the diversity of the training dataset, and thus to reduce the risk of overfitting the CNN. We applied several augmentations to each image before rescaling to a square. First, images were horizontally mirrored to mimic the left and right eye orientations of each image. Second, images were randomly cropped less than 10% on both height and width to mimic images with not well-centered macula. The augmented images were assigned the same labels as the corresponding original images. The purpose of image augmentation is to control for the overfitting problem by artificially creating training images, and these augmented images still preserve the key information of the image but are different from their original images. The proposed augmentation techniques were similar to the previous CNN study of fundus images ^17^. After augmentation, all fundus images were resized to squares encompassing the macula and rescaled to 224×224 pixels.

### Deep Learning Approach Combining Images and Genotypes

The first part of the model architecture is CNN used to extract features from fundus images (Figure 5). CNN is a special type of deep neural network that consists of many repeated processing layers that match the input image with successive convolutional filters to extract image features from low to high levels. A CNN is a member of deep neural networks that optimizes the weights of each layer using stochastic gradient descent via a backpropagation process. There can be millions of weights ^33^. There are a number of existing CNN architectures designed for image processing ^34^. In general, these architectures are similar. They are all comprised of sequentially convolutional and pooling layers. Each of the different architectures is best suited for specific problems. In this study, we used the Inception-v3 CNN architecture ^35^ to extract image features, which has been used for fundus images in several studies ^17,19,20,36^. Additionally, we used pre-trained weights as the initial values to train our network, which were trained for general image classification using the ImageNet database ^37^ that contains thousands of different objects and millions of images. This scale of data is usually unavailable in medical image classification studies.

**Figure 5.**
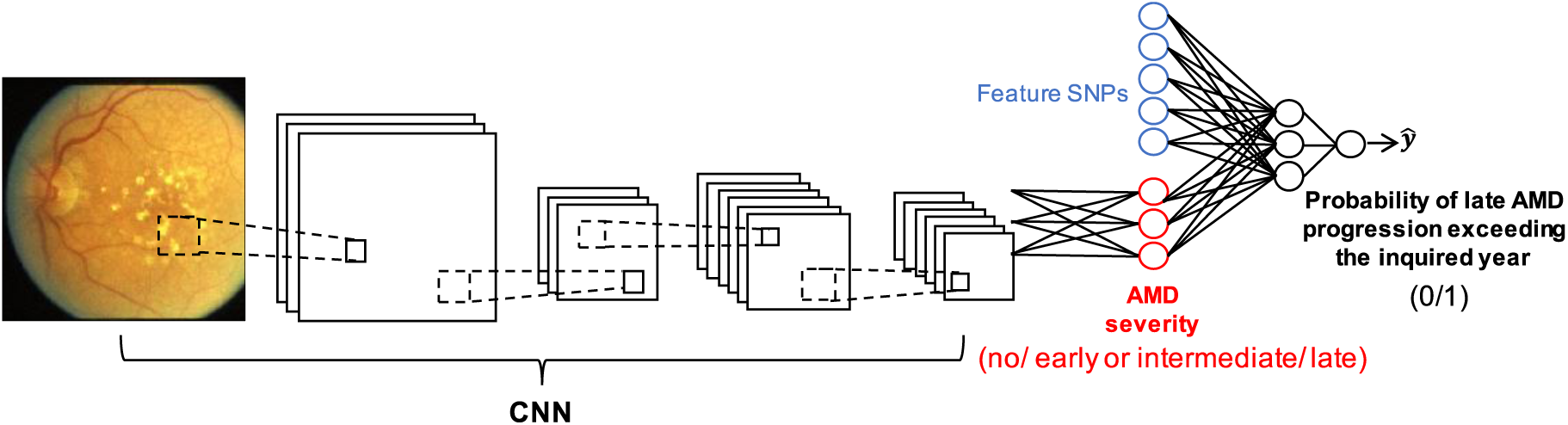
Convolutional neural network (CNN) of retinal fundus images along with feature SNPs and AMD severity for the prediction of late-AMD progression exceeding certain inquired years.

It was reported that the current AMD severity was the strongest predictor for the progression time to late AMD ^38^ and fundus images could be used to automatically grade AMD severity ^17,18,20^ with similar CNN architectures as presented here. Thus, after obtaining the output vector of the final convolutional layer, which contains all the information needed to understand the image, we fed these extracted image features to a fully connected layer to classify AMD severity (Figure 5). Furthermore, this severity viewed as the current AMD severity along with 52 independent genetic variants were fed to another fully connected layer to predict the time to late AMD development exceeding certain inquired years (Figure 5). During training, we used 3-level severity labels (e.g., no AMD when severity score is equal to 1; early or intermediate AMD when severity score is between 2 and 8; and late AMD when severity score is greater than or equal to 9). The network was built for any inquired year *k*, where *k* = 2, 3, 4, 5, 6, and 7. The detailed Inception-v3 CNN is shown in Supplementary Figure 7.

The aforementioned model could be simplified to sub-models. In total, we considered four models: (1) using fundus images taken at the current visit to predict whether the eye’s progression time to late AMD exceeds the inquired year; (2) using both fundus images and feature SNPs to predict whether the eye’s progression time to late AMD exceeds the inquired year; (3) using fundus images to predict whether the eye’s progression time to late AMD exceeds the inquired year as well as to classify the AMD severity at the current visit; (4) using both fundus images and feature SNPs to predict whether the eye’s progression time to late AMD exceeds the inquired year as well as to classify the AMD severity at the current visit.

Our deep learning network was implemented by using Keras with TensorFlow ^39^. During the training process, we first fixed the pre-trained weights in the CNN and updated the rest of the weights using an Adam optimizer with a learning rate of 0.001. The aforementioned 10-fold cross-validation was performed. Based on the average performance of the validation set, the best epoch was selected for testing set evaluation. For each fold, we trained the network for 20 epochs (loops of the entire training set) and selected the most suitable epoch for testing after 10-fold cross-validation. Furthermore, we set all weights as trainable and fine-tuned the network with a learning rate of 0.0001 and selected the best epoch out of 10 epochs also after 10-fold cross-validation. In this fine-tuning step, we selected a small total number of epochs to avoid updating the pre-trained CNN weights too much. All the training was conducted on a machine equipped with an EVGA GeForce RTX 2080 Ti 11Gb GPU and 128Gb available RAM.

### Performance Metrics

For the stepwise binary predictions, we calculated the AUC (area under the curve) of Receiver Operator Characteristic (ROC) curves as the primary performance metric. Besides, Brier score ^40^ that is the squared error of a probabilistic prediction was used as another metric, and the lower the Brier score the better the model predicts. The useful benchmark values for the Brier score are 33%, which corresponds to predicting the risk by a random number drawn from a Uniform distribution between 0 and 1. For the 3-level AMD severity classification, no AMD, early or intermediate AMD and late AMD were treated as levels 0, 1 and 2 (see supplementary text for details).

### Visualizing Model Attention

To help understand what image features were learned and make the “black box” deep learning model more transparent, we generated saliency maps ^41^ to highlight the regions that most contribute to the predicted values from the output layer for all trained models. The saliency is computed by the gradient of output value with respect to the input image. In other words, it could detect how a small change in the input image changes the output value. If these gradients have the same shape as the region of interest, it indicates that the attention of learning is on the right region. Thus, these gradients could be used to highlight input regions that result in the most change in the output prediction. The saliency map method only works when images are the only input. When both images and SNPs were inputs, we used LIME (Local Interpretable Model-Agnostic Explanations) ^42^ instead, which attempts to understand the model by perturbing the input data and understanding how the predictions change. Specifically, we extracted the network inside the red rectangle in Supplementary Figure 7b. After processing images through Inception-v3, an average pooling was used to generate a vector of length 2,048 (i.e., V1, V2 … V2048), which summarized the image information. Then, this vector was concatenated with the SNP vector of 52 SNPs. Therefore, the input for this sub-network is a vector of 2,100 elements, which is what we perturbed in our LIME analyses.

### Data availability

All the phenotype data and fundus images of AREDS participants required are available from dbGap (accession: phs000001.v3.p1). The genotype data on AREDS subjects has been reported earlier ^9^ and is available from dbGap (accession phs001039.v1.p1). The UK Biobank test dataset was obtained from UK Biobank (application number 43252).

### Code availability

The prediction models with Python implementation and a detailed tutorial are available at https://github.com/QiYanPitt/AMDprogressCNN and a web-based graphical user interface is also available at http://www.pitt.edu/~qiy17/amdprediction.html.

## Data Availability

All the phenotype data and fundus images of AREDS participants required are available from dbGap (accession: phs000001.v3.p1). The genotype data on AREDS subjects has been reported earlier 11 and is available from dbGap (accession phs001039.v1.p1). The UK Biobank test dataset was obtained from UK Biobank (application number 43252).

https://www.ncbi.nlm.nih.gov/projects/gap/cgi-bin/study.cgi?study_id=phs000001.v3.p1

https://www.ncbi.nlm.nih.gov/projects/gap/cgi-bin/study.cgi?study_id=phs001039.v1.p1

## Competing interests

The authors declare no competing financial and non-financial interest.

## ACKNOWLEDGEMENTS

H.H. was partially supported by NSF IIS 1852606 and IIS 1837956. A.S. was supported by NEI Intramural Research Program grant # ZIAEY000546. The independent validation has been conducted using the UK Biobank Resource under Application Number 43252.

## AUTHOR CONTRIBUTIONS

Conception and Project Supervision: Q.Y., Y.D., W.C.; Data Processing and Analysis: Q.Y.; Study design: Q.Y., H.X.; Data Interpretation: Q.Y., A.S., E.Y.C.; Writing: Q.Y. and D.E.W.; Critical Review of Manuscript: D.E.W., Y.D., W.C., H.H., A.S. and E.Y.C.

## Tables

**Supplementary Table 1.**
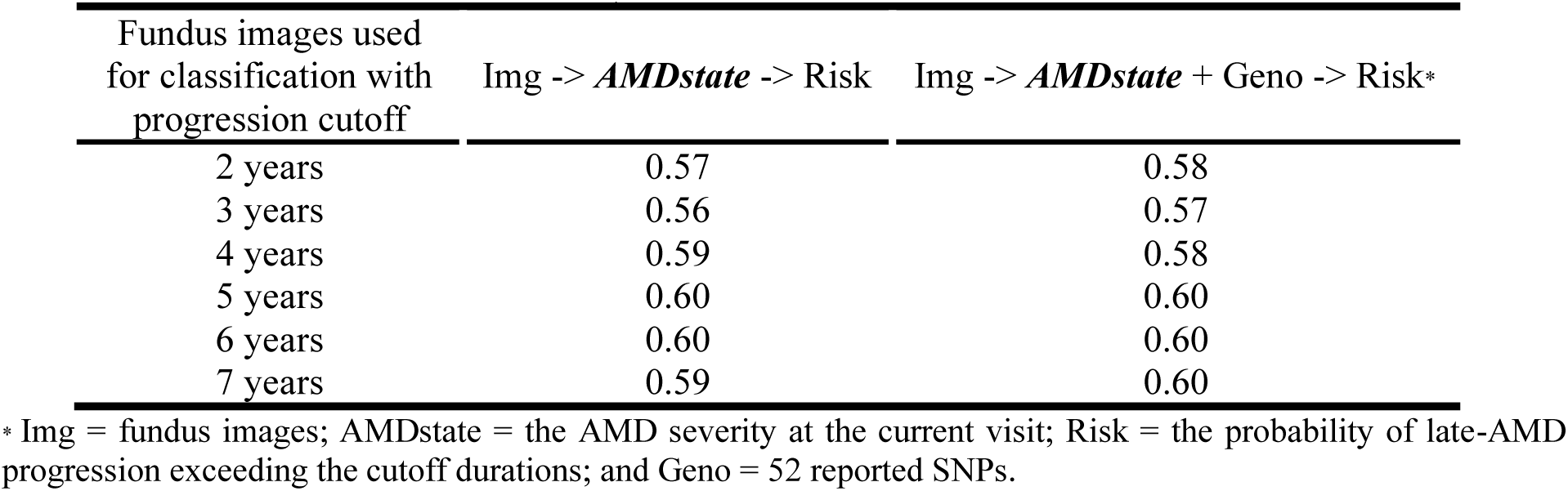
The 3-level AMD severity classification accuracy (no AMD, early or intermediate AMD and late AMD)

| Fundus images used<br>for classification with<br>progression cutoff | Img -> *AMDstate* -> Risk | Img -> *AMDstate* + Geno -> Risk* |
| --- | --- | --- |
| 2 years | 0.57 | 0.58 |
| 3 years | 0.56 | 0.57 |
| 4 years | 0.59 | 0.58 |
| 5 years | 0.60 | 0.60 |
| 6 years | 0.60 | 0.60 |
| 7 years | 0.59 | 0.60 |

\* Img = fundus images; AMDstate = the AMD severity at the current visit; Risk = the probability of late-AMD progression exceeding the cutoff durations; and Geno = 52 reported SNPs.

**Supplementary Table 2.**
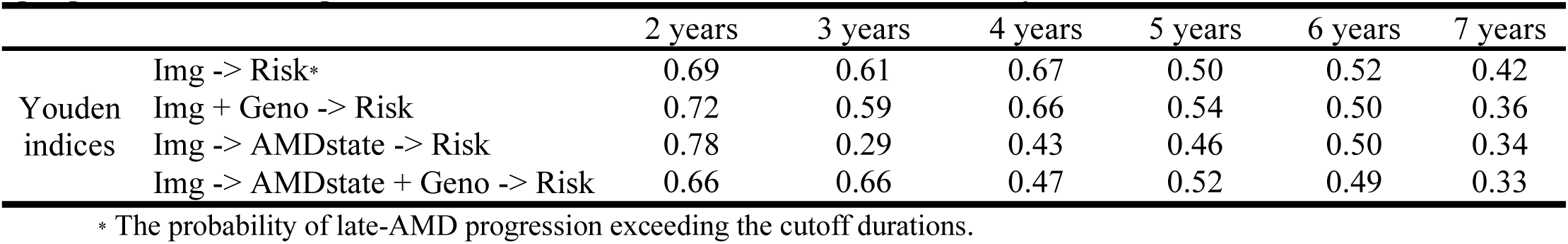
Youden indices for dichotomizing the prediction of probability of late-AMD progression exceeding the cutoff durations for four models with cutoff years

|  |  | 2 years | 3 years | 4 years | 5 years | 6 years | 7 years |
| --- | --- | --- | --- | --- | --- | --- | --- |
| Youden indices | Img -> Risk* | 0.69 | 0.61 | 0.67 | 0.50 | 0.52 | 0.42 |
|  | Img + Geno -> Risk | 0.72 | 0.59 | 0.66 | 0.54 | 0.50 | 0.36 |
|  | Img -> AMDstate -> Risk | 0.78 | 0.29 | 0.43 | 0.46 | 0.50 | 0.34 |
|  | Img -> AMDstate + Geno -> Risk | 0.66 | 0.66 | 0.47 | 0.52 | 0.49 | 0.33 |

\* The probability of late-AMD progression exceeding the cutoff durations.

### The 3-level AMD severity classification

No AMD (level 0) was coded as 00, early or intermediate AMD (level 1) was coded as 10 and late AMD (level 2) was coded as 11 (Supplementary Table 3) so that the order of classes could be learned. The performance metric used was accuracy of predicted labels. The predicted labels were determined based on the highest predicted label probability. For the predicted probability of each label:

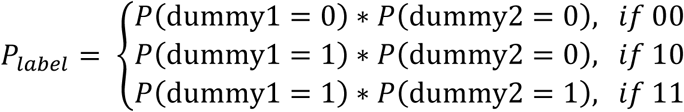

where *P*(dummy1 = 0)is the predicted probability of 0 for dummy1 and *P*(dummy1 = 1)is the predicted probability of 1 for dummy1. It is analogous to dummy2. The label was assigned by the highest probability. For example, if *P*(dummy1 = 1)\**P*(dummy2= 1)had the highest probability, the predicted label would be late AMD. Then, the accuracy was the number of correct predicted labels divided by the sample size. Note that the random accuracy is one out of three, 0.33.

**Supplementary Table 3.** Dummy variables for eye AMD severity at the current visit

|  | dummy1 | dummy2 |
| --- | --- | --- |
| no AMD | 0 | 0 |
| intermediate AMD | 1 | 0 |
| late AMD | 1 | 1 |

**Supplementary Figure 1.**
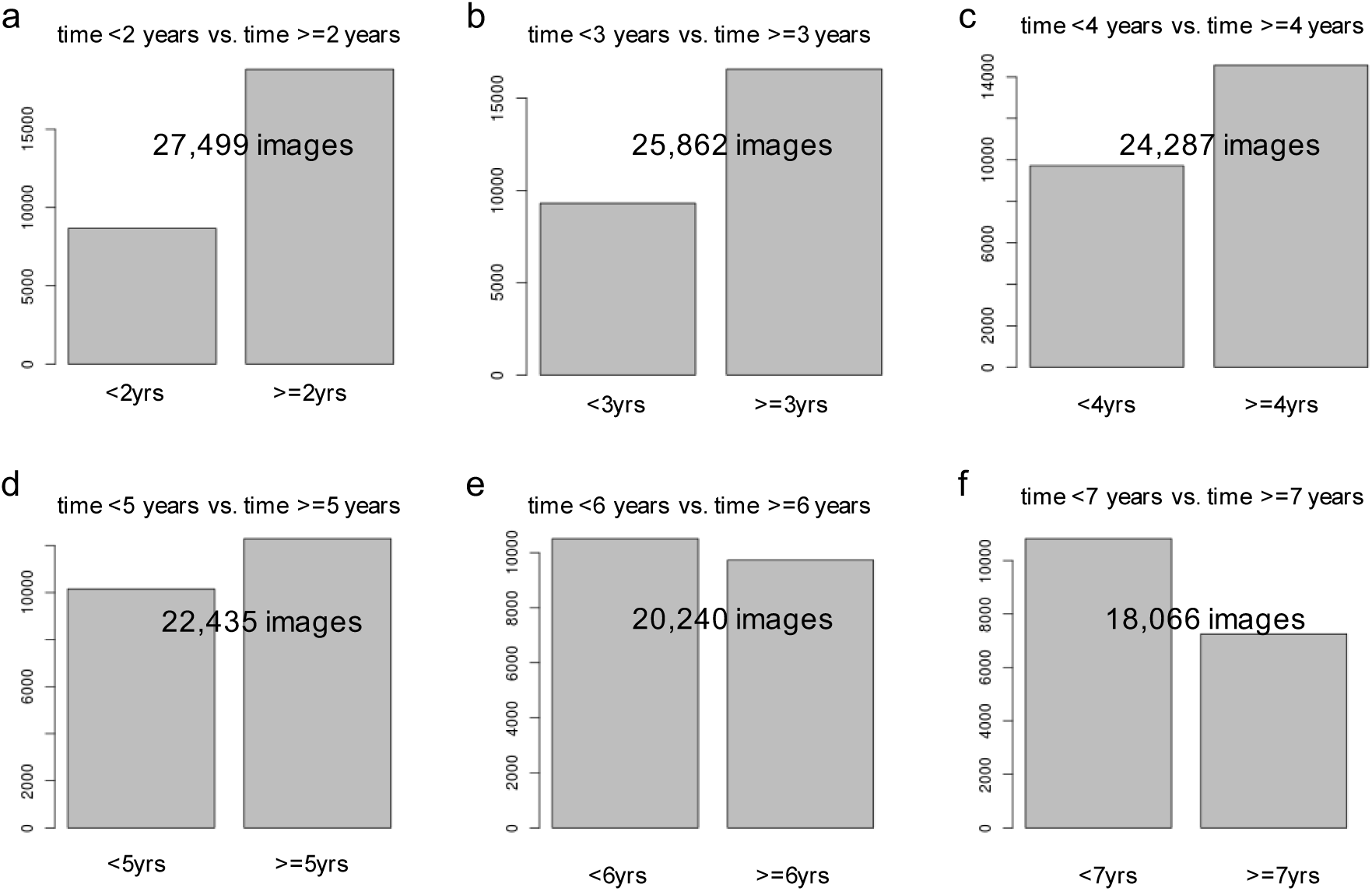
The number of useable fundus images for prediction before and after the cutoff years (2, 3, 4, 5, 6 and 7 years)

**Supplementary Figure 2.**
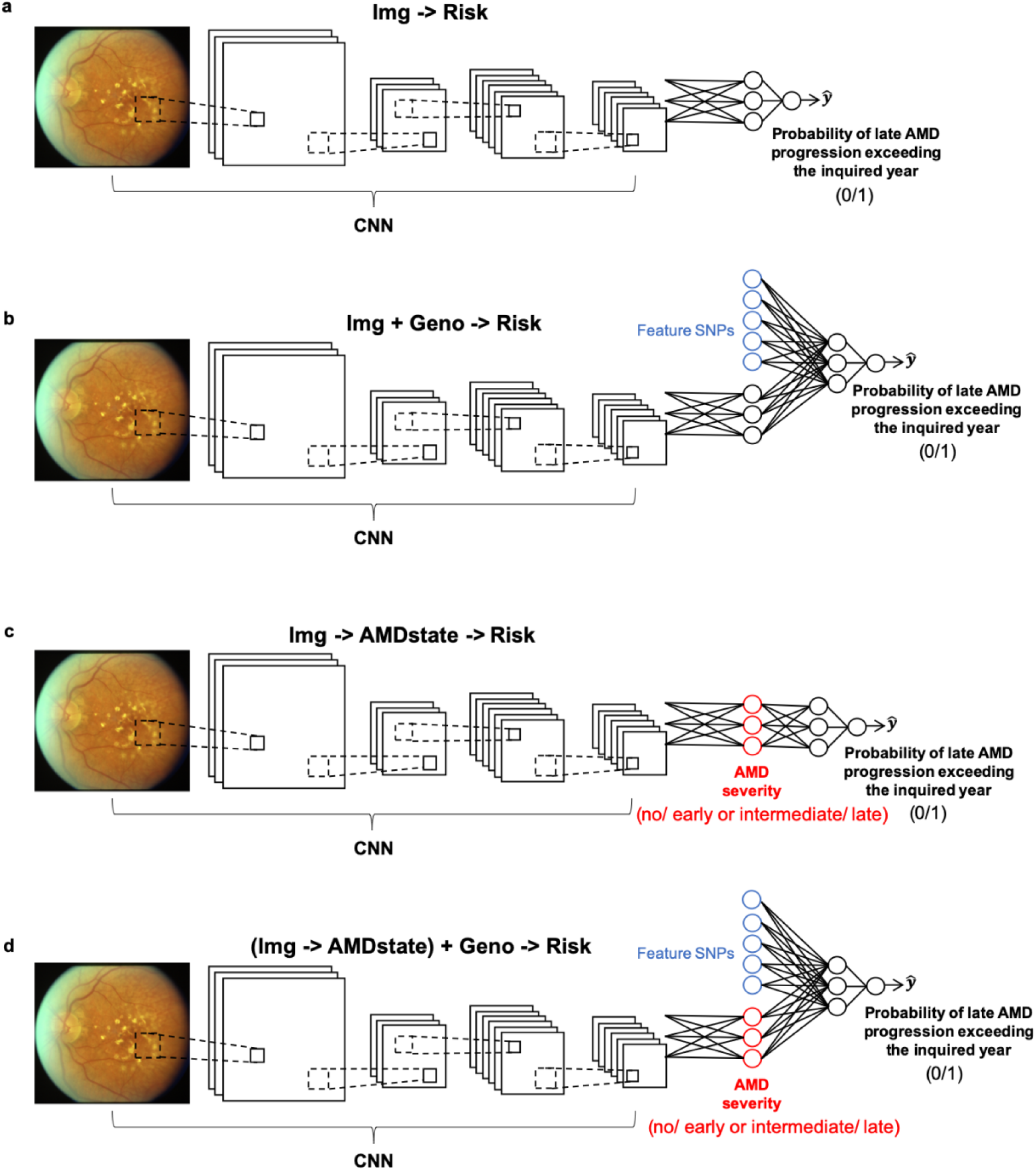
Four models of convolutional neural network (CNN) for AMD progression time prediction. (a) fundus images predicting late-AMD progression exceeding the cutoff durations; (b) fundus images + genotypes predicting late-AMD progression exceeding the cutoff durations; (c) fundus images both classifying current AMD severity and predicting late-AMD progression exceeding the cutoff durations; and (d) fundus images + genotypes both classifying current AMD severity and predicting late-AMD progression exceeding the cutoff durations.

**Supplementary Figure 3.**
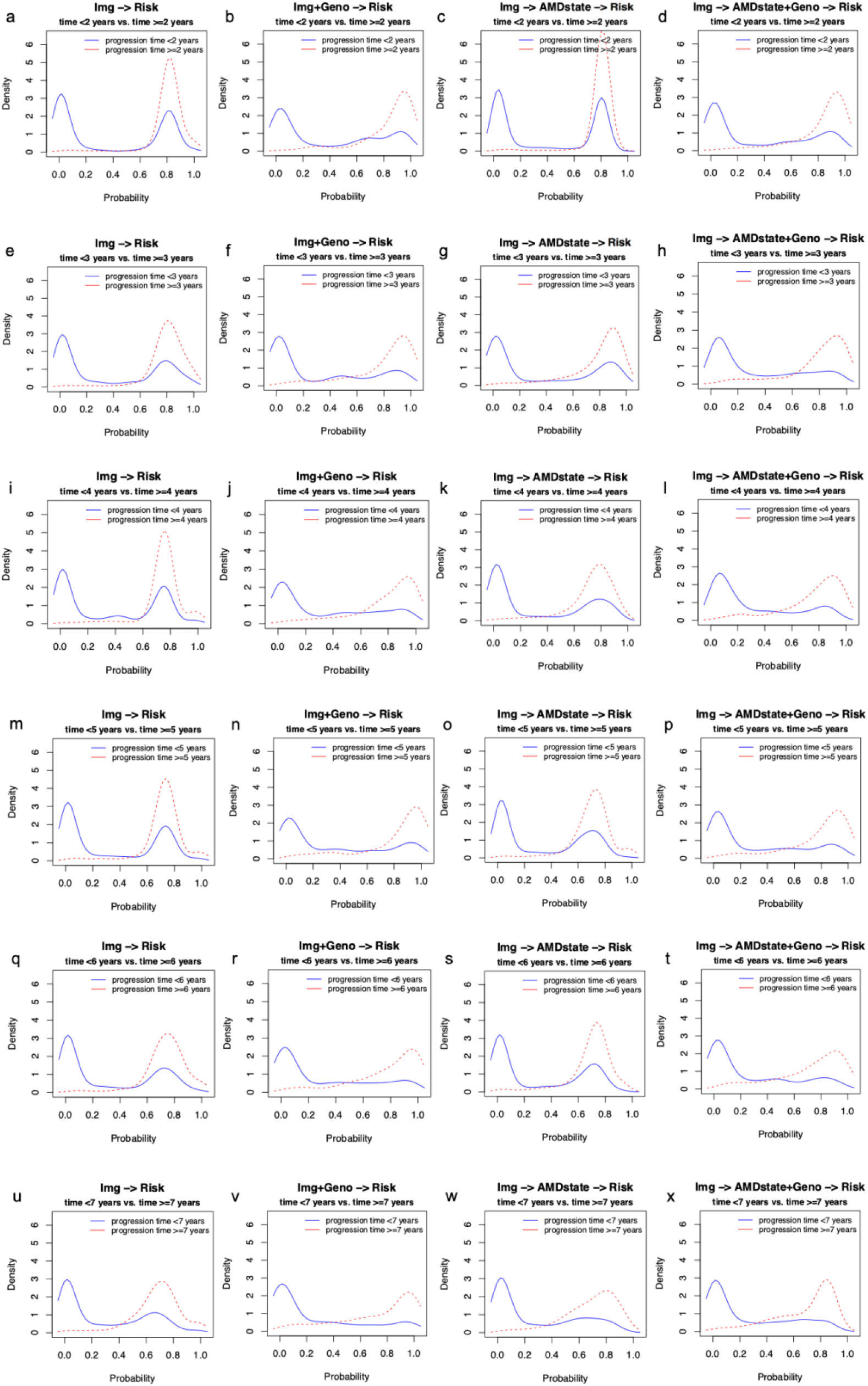
Density curves of the predicted probability of having late AMD progression time before or after each of the six cutoff years for four models. Each column is one model and each row is one cutoff year.

**Supplementary Figure 4.**
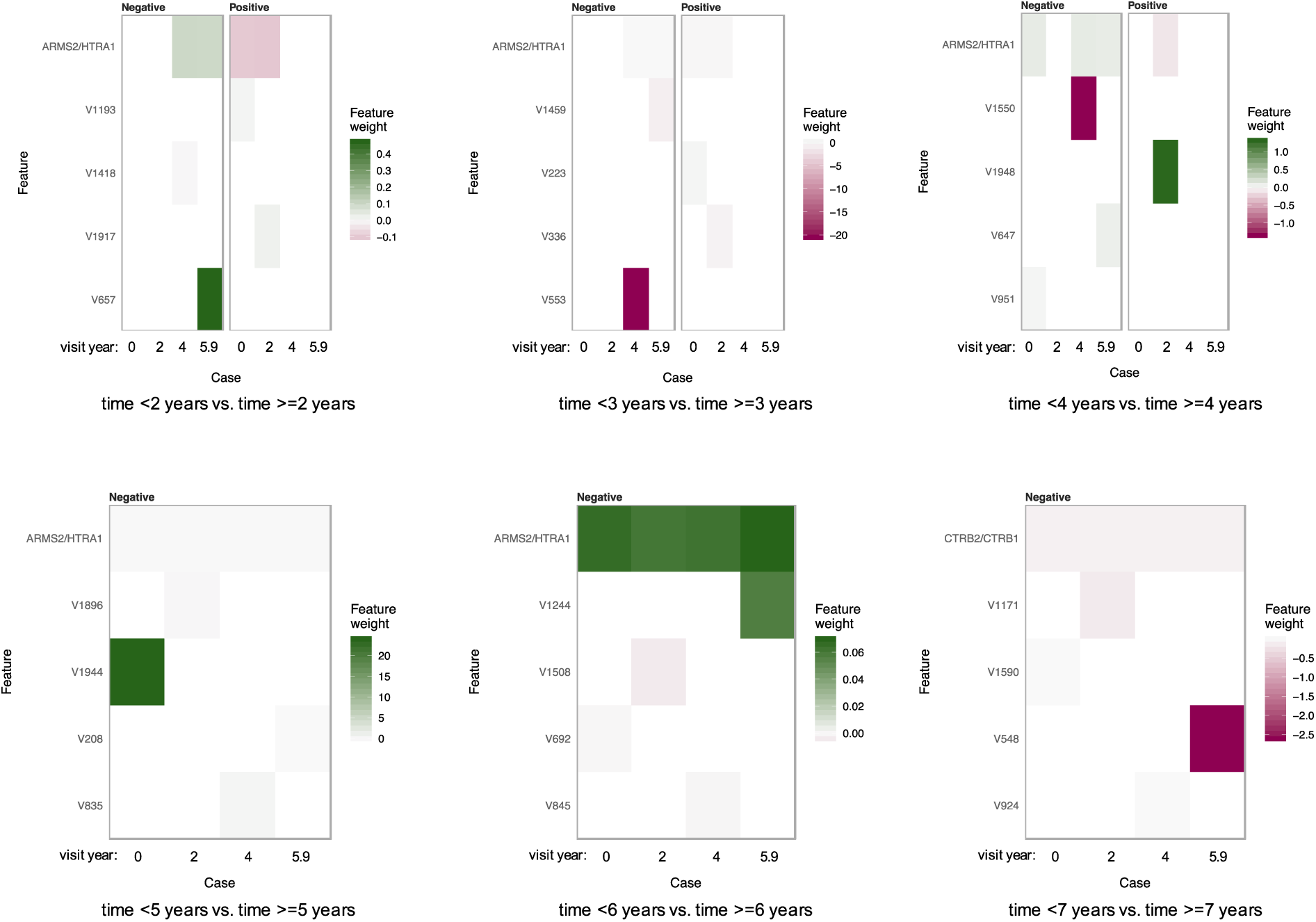
Feature importance heatmaps from LIME for the sub-network using fundus images along with genotypes (Supplementary Figure 7b inside the red rectangle) to predict late AMD progression time for the same left eye of Subject #1 in Figure 2. This subject progressed to late AMD after 4.8 years of follow-up. The first two variables contributing the most to the predictions for each visit year are shown. Note that with each visit year, we selected the two variables with the most extreme feature weights for display here. The V#s indicates the summarized image information (i.e., V1, V2 … V2048).

**Supplementary Figure 5.**
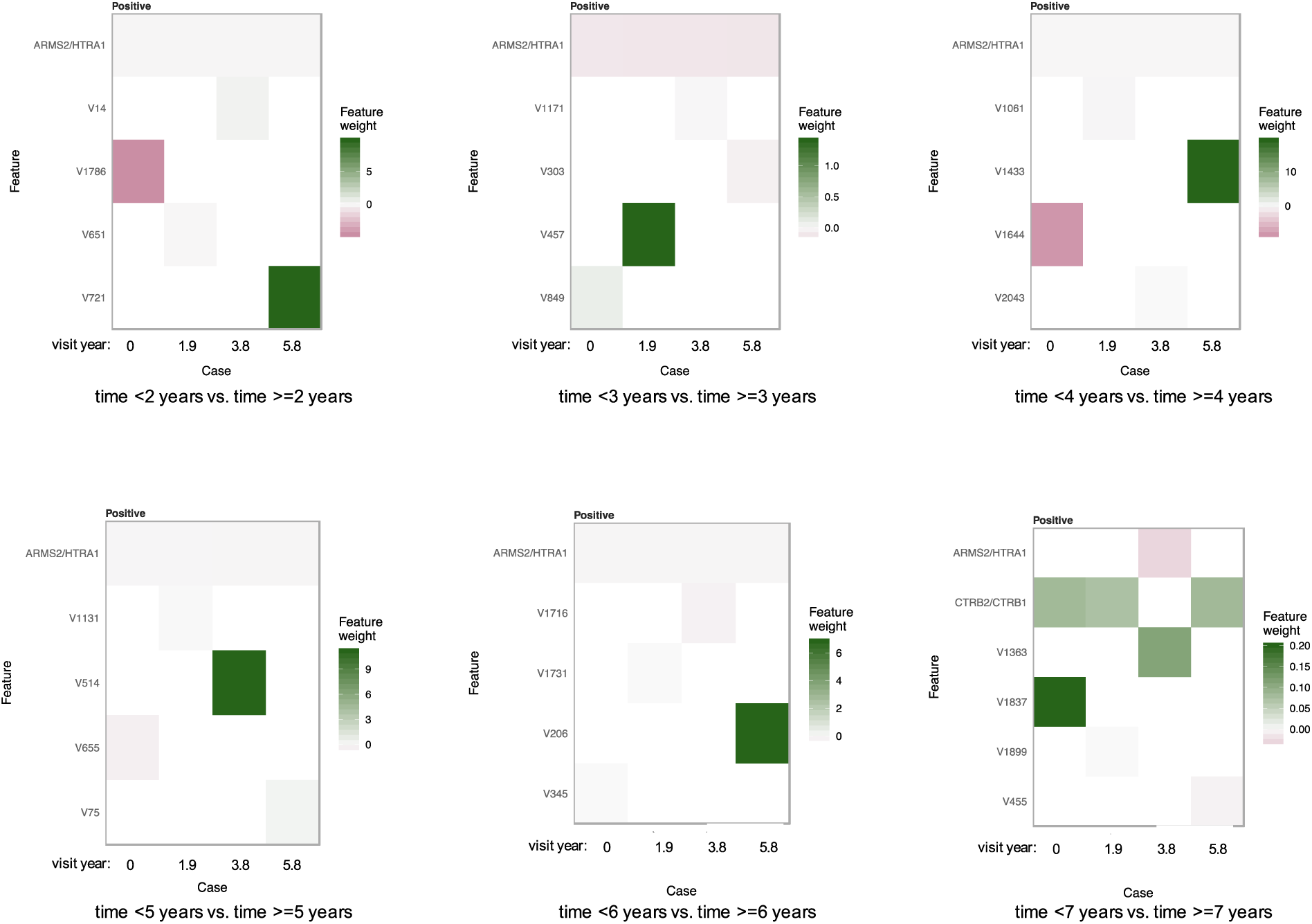
Feature importance heatmaps from LIME for the sub-network using fundus images along with genotypes (Supplementary Figure 7b inside the red rectangle) to predict late AMD progression time for the same left eye of Subject #2 in Figure 3. This subject was censored after 11.1 years of follow-up. The first two variables contributing the most to the predictions for each visit year are shown. Note that with each visit year, we selected the two variables with the most extreme feature weights for display here. The V#s indicates the summarized image information (i.e., V1, V2 … V2048).

**Supplementary Figure 6.**
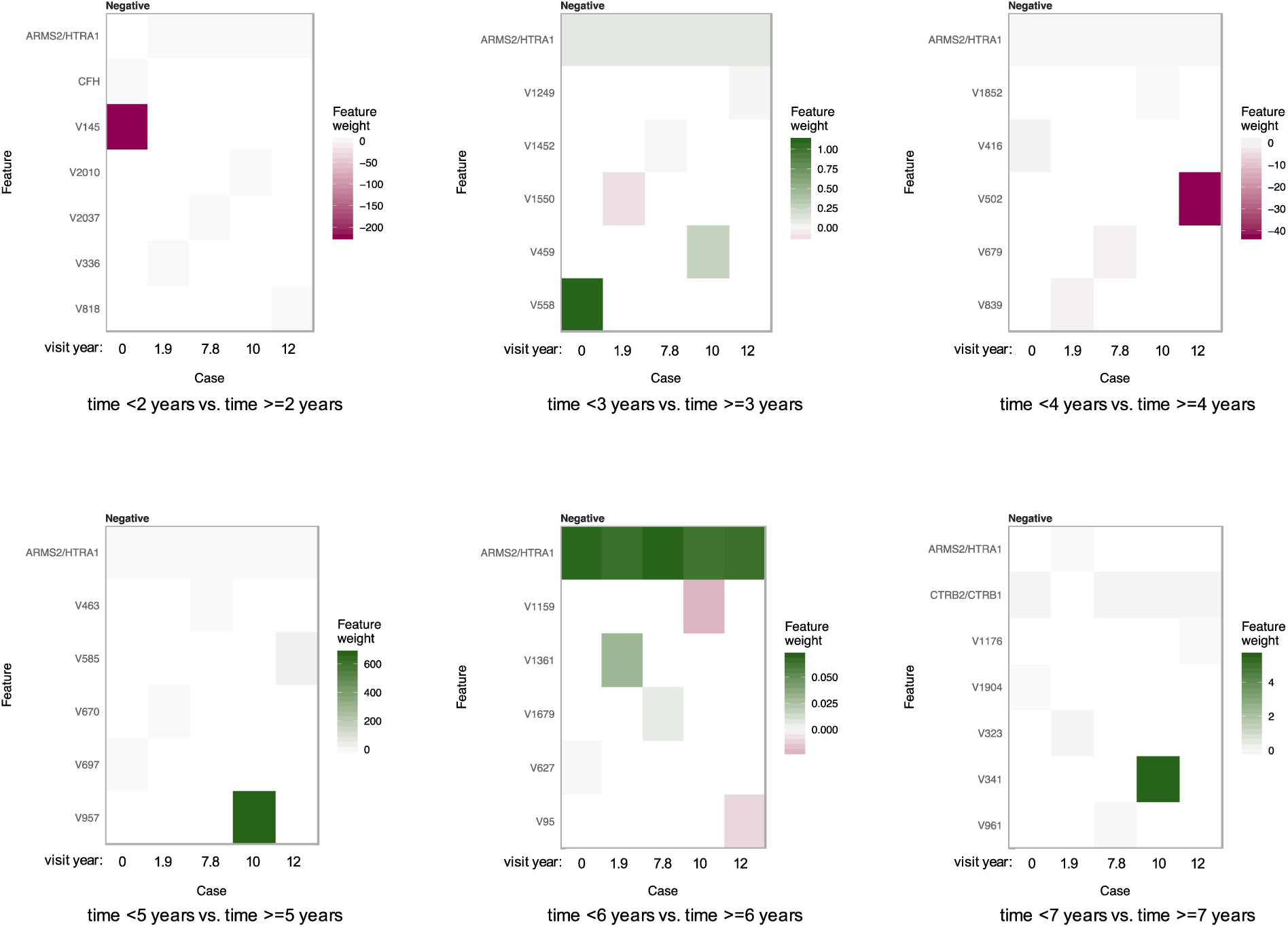
Feature importance heatmaps from LIME for the sub-network using fundus images along with genotypes (Supplementary Figure 7b inside the red rectangle) to predict late AMD progression time for the same left eye of Subject #3 in Figure 4. This subject developed late AMD before enrollment. The first two variables contributing the most to the predictions for each visit year are shown. Note that with each visit year, we selected the two variables with the most extreme feature weights for display here. The V#s indicates the summarized image information (i.e., V1, V2 … V2048).

**Supplementary Figure 7.**
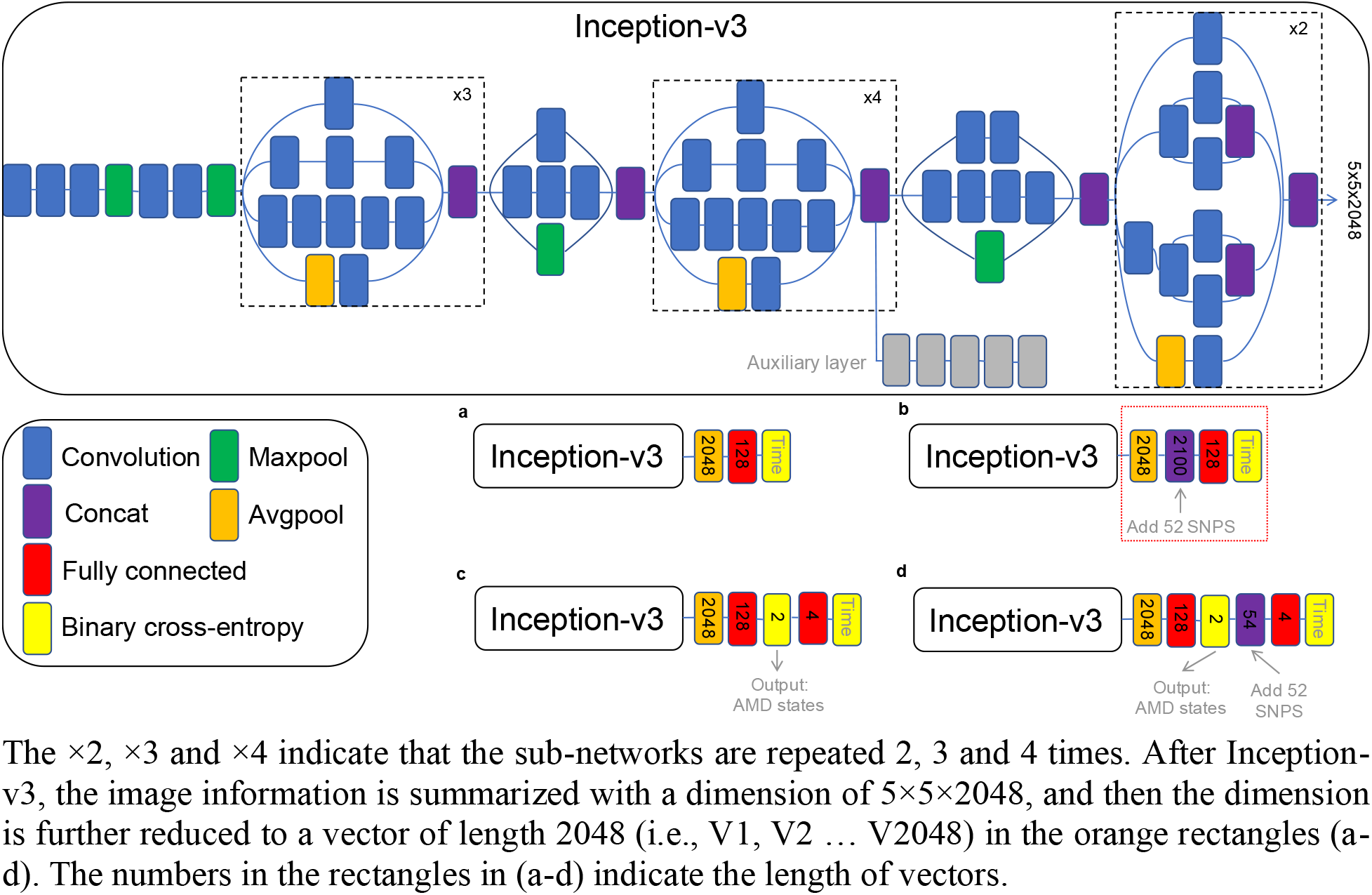
Detailed Inception-v3 convolutional neural network (CNN) for AMD progression time prediction. (a) fundus images predicting late-AMD progression exceeding the cutoff durations; (b) fundus images + genotypes predicting late-AMD progression exceeding the cutoff durations; (c) fundus images both classifying current AMD severity and predicting late-AMD progression exceeding the cutoff durations; and (d) fundus images + genotypes both classifying current AMD severity and predicting late-AMD progression exceeding the cutoff durations. Note that the auxiliary layer was not included in this Keras version Inception-v3 network.

**Supplementary Figure 8.**
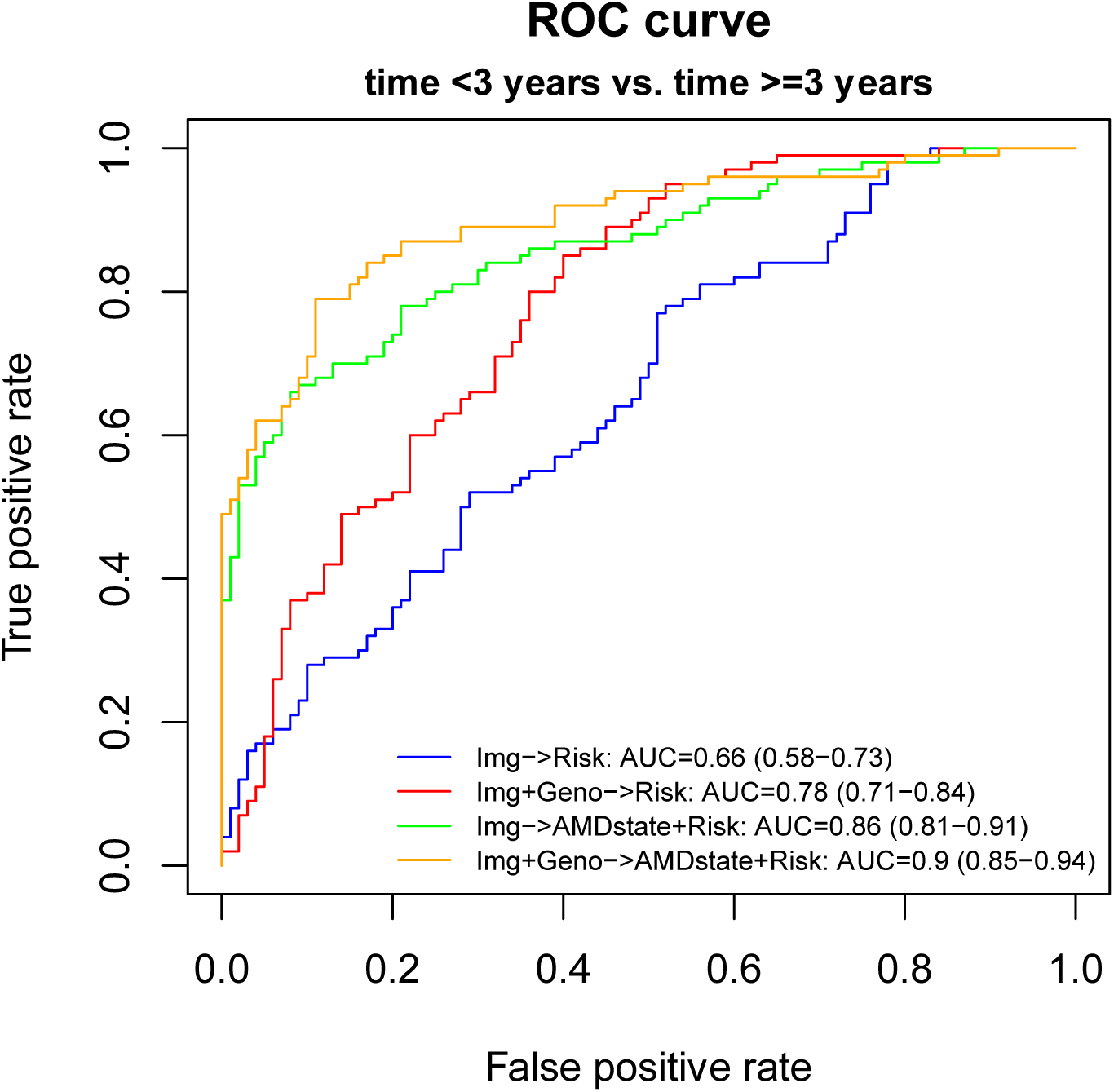
ROC curves of the classification of late AMD progression exceeding the cutoff duration of 3 years for four models using 200 Caucasians from UK Biobank. The four models are (1) fundus images predicting late-AMD progression exceeding 3 years; (2) fundus images + genotypes predicting late-AMD progression exceeding 3 years; (3) fundus images both classifying current AMD severity and predicting late-AMD progression exceeding 3 years; and (4) fundus images + genotypes both classifying current AMD severity and predicting late-AMD progression exceeding 3 years.

**Supplementary Figure 9.**
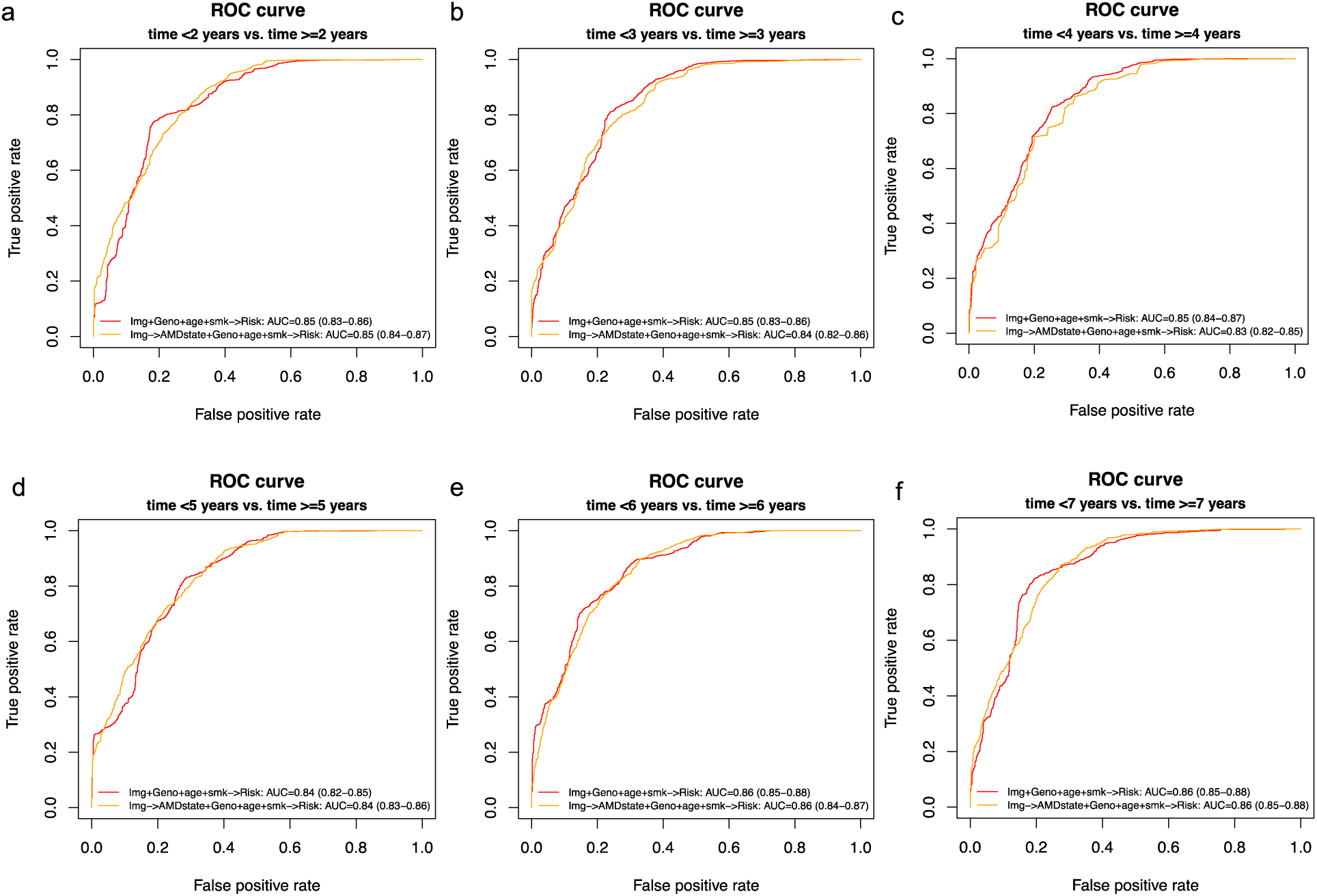
ROC curves of the prediction of late AMD progression time exceeding the inquired years for four models. **The two models are** (1) fundus images + genotypes + age + smoking status predicting late-AMD progression exceeding the inquired years; and (2) fundus images + genotypes + age + smoking status both classifying current AMD severity and predicting late-AMD progression exceeding the inquired years. (a-f) inquired years from 2 to 7.

**Supplementary Figure 10.**
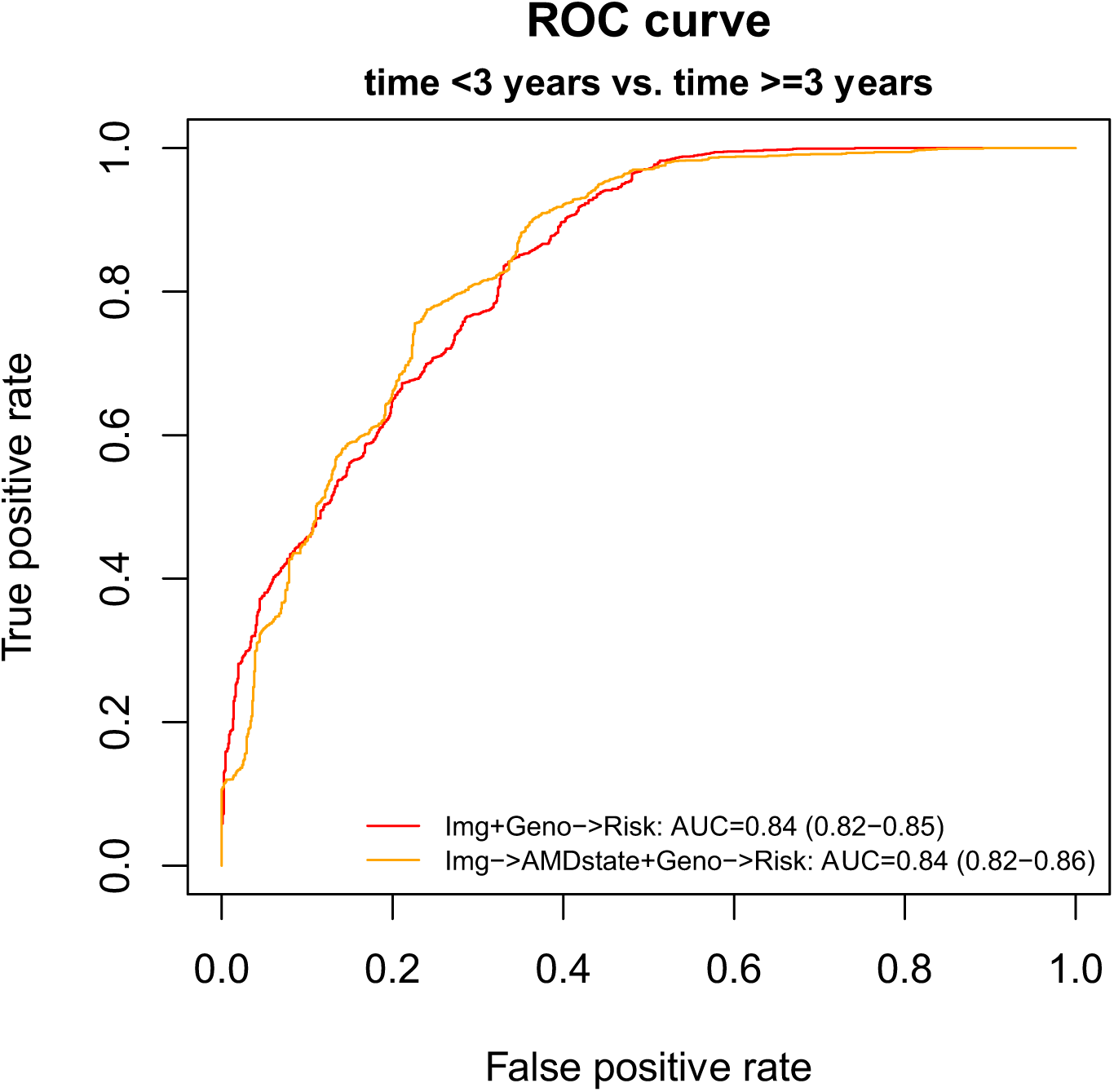
ROC curves of the classification of late AMD progression exceeding the cutoff duration of 3 years using fundus images + 1,057 SNPs. The two models are (1) fundus images + genotypes predicting late-AMD progression exceeding 3 years; and (2) fundus images + genotypes both classifying current AMD severity and predicting late-AMD progression exceeding 3 years.

**Supplementary Figure 11.**
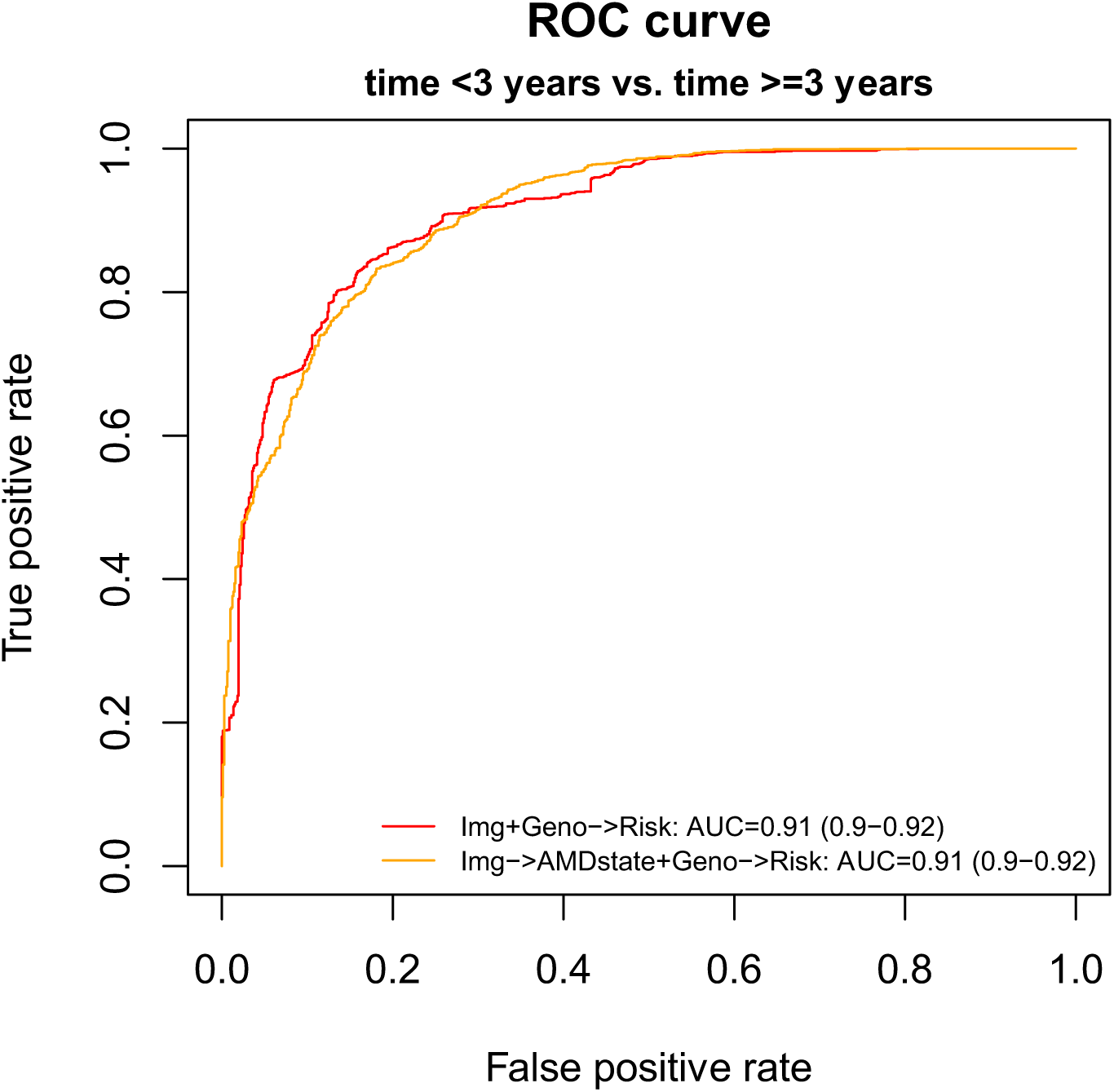
ROC curves of the classification of late AMD progression exceeding the cutoff duration of 3 years using fundus images + 52 SNPs + the other eye’s current severity. The two models are (1) fundus images + genotypes + the other eye’s current severity predicting late-AMD progression exceeding 3 years; and (2) fundus images + genotypes + the other eye’s current severity both classifying current AMD severity and predicting late-AMD progression exceeding 3 years.

## Notes

### Competing Interest Statement

The authors have declared no competing interest.

### Funding Statement

No external funding was received

### Author Declarations

All relevant ethical guidelines have been followed and any necessary IRB and/or ethics committee approvals have been obtained.

Any clinical trials involved have been registered with an ICMJE-approved registry such as ClinicalTrials.gov and the trial ID is included in the manuscript.

